# Projected COVID-19 epidemic in the United States in the context of the effectiveness of a potential vaccine and implications for social distancing and face mask use

**DOI:** 10.1101/2020.10.28.20221234

**Authors:** Mingwang Shen, Jian Zu, Christopher K. Fairley, José A. Pagán, Li An, Zhanwei Du, Yuming Guo, Libin Rong, Yanni Xiao, Guihua Zhuang, Yan Li, Lei Zhang

## Abstract

**Background:** Multiple candidates of COVID-19 vaccines have entered Phase III clinical trials in the United States (US). There is growing optimism that social distancing restrictions and face mask requirements could be eased with widespread vaccine adoption soon.

**Methods:** We developed a dynamic compartmental model of COVID-19 transmission for the four most severely affected states (New York, Texas, Florida, and California). We evaluated the vaccine effectiveness and coverage required to suppress the COVID-19 epidemic in scenarios when social contact was to return to pre-pandemic levels and face mask use was reduced. Daily and cumulative COVID-19 infection and death cases were obtained from the Johns Hopkins University Coronavirus resource center and used for model calibration.

**Results:** Without a vaccine, the spread of COVID-19 could be suppressed in these states by maintaining strict social distancing measures and face mask use levels. But relaxing social distancing restrictions to the pre-pandemic level without changing the current face mask use would lead to a new COVID-19 outbreak, resulting in 0.8-4 million infections and 15,000-240,000 deaths across these four states over the next 12 months. In this scenario, introducing a vaccine would partially offset this negative impact even if the vaccine effectiveness and coverage are relatively low. However, if face mask use is reduced by 50%, a vaccine that is only 50% effective (weak vaccine) would require coverage of 55-94% to suppress the epidemic in these states. A vaccine that is 80% effective (moderate vaccine) would only require 32-57% coverage to suppress the epidemic. In contrast, if face mask usage stops completely, a weak vaccine would not suppress the epidemic, and further major outbreaks would occur. A moderate vaccine with coverage of 48-78% or a strong vaccine (100% effective) with coverage of 33-58% would be required to suppress the epidemic. Delaying vaccination rollout for 1-2 months would not substantially alter the epidemic trend if the current interventions are maintained.

**Conclusions:** The degree to which the US population can relax social distancing restrictions and face mask use will depend greatly on the effectiveness and coverage of a potential COVID-19 vaccine if future epidemics are to be prevented. Only a highly effective vaccine will enable the US population to return to life as it was before the pandemic.

## INTRODUCTION

The coronavirus disease (COVID-19) is resulting in enormous health and economic losses in the United States (US).^1–4^ As of 20th October 2020, there are more than 8 million cases of COVID-19 and more than 220,000 deaths in the US.^5^ The cooler weather in the US is seeing evidence of second waves of infection occurring in many US states.^5^ Some US politicians are suggesting that an effective vaccine would mean that Americans could return to normal life so that citizens would no longer need to socially distance or wear face masks, and the economy can be fully revived.

However, the degree to which these restrictions could be eased will be dependent on the effectiveness of the potential COVID-19 vaccines which is currently unknown.^6–8^ To allow careful planning about what restrictions may need to be continued, research is urgently needed to project how the effectiveness of a potential vaccine may affect the trajectory of the COVID-19 pandemic in the US. It is also important to determine how the current non-pharmaceutical interventions can be integrated into an overall COVID-19 control strategy that includes vaccines of different effectiveness.^9^ There are three key questions that need to be addressed to plan an effective control strategy once an effective vaccine becomes available. These are: (1) How effective would the vaccine need to be to suppress the pandemic? (2) What proportion of the population would need to be vaccinated? and (3) What levels of social distancing measures and face mask use would need to be maintained in the context of different values of vaccine effectiveness and coverage?

To address these questions, we developed dynamic simulation models of COVID-19 for the four hardest-hit states in the US (New York, Texas, Florida, and California). The models were developed for each state because each state has different COVID-19 policies for social distancing measures and mask use. Each of the state-level models was calibrated based on the most recent COVID-19 data of that state. The models were then used to project the averted COVID-19 cases and deaths at different levels of vaccine effectiveness and coverage for the four states. Given that the proportion of people who wear face masks is different across the four states, we further examined how face mask use would influence the effect of a potential vaccine in controlling the pandemic. Findings from this study provide timely information that can be used by policymakers to plan for the potential release of a COVID-19 vaccine and understand its effect across different regions in the US under different social distancing and face mask use scenarios.

## METHODS

### Data sources

We obtained COVID-19 data for New York, Texas, Florida, and California from the Johns Hopkins University Coronavirus Resource Center.^5^ The data included the number of daily and cumulative confirmed cases and deaths from 26th January to 15th September 2020. These data were used to calibrate the model for each state.

### Model formulation and assumptions

We developed a dynamic compartmental model to capture the transmission of COVID-19 in each state. **Figure 1** shows the structure of our model. The population in each state was divided into ten compartments (susceptible individuals (S), vaccinated individuals (V), latent infections (E), asymptomatic infections (A, infected individuals without symptoms), undiagnosed infections with mild/moderate (I_1_) and severe/critical symptoms (I_2_), diagnosed infections with mild/moderate (T_1_) or severe/critical symptoms (T_2_), and recovered (R) and deceased (D) cases). Susceptible and vaccinated individuals could be infected by contacts with latent, asymptomatic, and undiagnosed infections with mild/moderate and severe/critical symptoms in public settings (e.g., public transportation, supermarkets, workplaces, etc.) and households (home or other private settings) with a probability *Λ* and Λ_*V*_ (the force of infection), respectively (detailed descriptions of *Λ* and Λ_*V*_ are provided in the Supplementary Appendix). Our model did not consider population mobility in these states.

**Figure 1.**
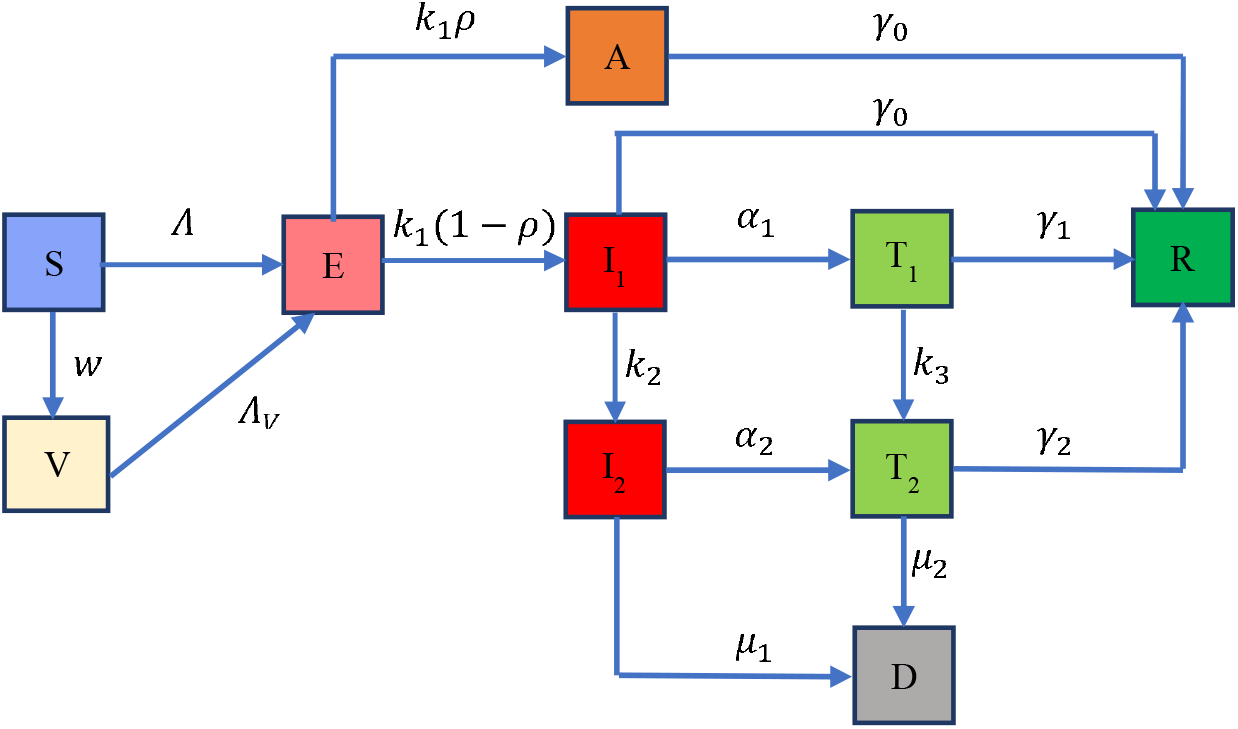
Flow chart of COVID-19 transmission model. The total population is divided into ten compartments (susceptible individuals (S), vaccinated individuals (V), latent infections (E), asymptomatic infections (A), undiagnosed infections with mild/moderate (I_1_) and severe/critical symptoms (I_2_), diagnosed infections with mild/moderate (T_1_) and severe/critical symptoms (T_2_), and recovered (R) and deceased (D) cases). The force of infection for susceptible and vaccinated individuals are denoted as *Λ* and Λ_*V*_, which involves two transmission patterns: public settings (e.g., public transportation, supermarkets, offices, etc.) and households. The model includes three control measures: handwashing, social distancing and face mask use. More details on *Λ* and Λ_*V*_ are provided in the **Appendix**. The average incubation period is 1/*k*_1,_ and the probability that an individual is asymptomatic is *ρ*. Infectious individuals with mild/moderate and severe/critical symptoms are diagnosed and treated at the rates *α*_l_ and *α*_2_, respectively. We assume these diagnosed individuals are isolated strictly and could not further infect others. Undiagnosed and diagnosed mild/moderate cases progress to the severe/critical stage at the rates *k*_2_and *k*_3_, respectively. Asymptomatic infections and undiagnosed mild/moderate cases are assumed to recover naturally at the rate *γ*_0_. Diagnosed mild/moderate and severe/critical cases will recover at the rates *γ*_l_ and *γ*_2_, respectively. Undiagnosed and diagnosed severe/critical cases will die due to the disease at the rates *μ*_l_ and *μ*_2_, respectively. The vaccination rate is denoted as *w*.

Latent individuals could progress to the infectious compartments with mild/moderate symptoms or asymptomatic compartments at a rate *k*_1_. The probability that an individual is asymptomatic after an infection is *ρ*. Infectious individuals with mild/moderate and severe/critical symptoms are diagnosed and treated at the rates *α*_l_ and *α*_2_, respectively. We assumed that diagnosed individuals were isolated and could not infect others. Undiagnosed and diagnosed mild/moderate cases could progress to the severe/critical stages at the rates *k*_2_ and *k*_3_, respectively. Asymptomatic infections and undiagnosed mild/moderate cases were assumed to recover naturally at the rate *γ*_0_. Diagnosed mild/moderate and severe/critical cases could recover at the rates *γ*_l_ and *γ*_2_, respectively. Undiagnosed and diagnosed severe/critical cases could die due to the disease at the rates *μ*_l_ and *μ*_2_, respectively.

We assumed that the face mask effectiveness to prevent infection is 85% (95% CI, 66%-93%) based on a meta-analysis on the effectiveness of face masks for COVID-19.^10^ We obtained data on the proportion of people who always wear a face mask at the county level from The New York Times (based on about 250,000 interviews conducted by Dynata from 2nd July to 14th July 2020.^11^ We then estimated state-level face mask use by combining county-level data. The proportions of people who always wear a face mask in New York, Texas, Florida, and California were estimated to be 76.6%, 71.7%, 58.7%, and 76.6%, respectively.

We denoted the vaccination rate as *w* and the effectiveness of the vaccine against the acquisition of infection as ε_*V*_. That is, the probability of being infected for a vaccinated individual was 1-ε_*V*_ of that for an unvaccinated individual when the vaccine is available. There is no COVID-19 vaccine data publicly available in the US right now; as such, we varied the vaccine effectiveness ε_*V*_ from 0 to 100% and assumed the vaccine coverage rate (V/(V+S)) changed from 0 to 100% by varying the vaccination rate *w*.

### Model calibration

We estimated some of the model parameters using a nonlinear least-squares method (see Supplementary Appendix) and calibrated the model using data on daily and cumulative confirmed infections and deaths. **Figure 2** shows the model calibration results for all the four states. The other model parameters were estimated from the literature (see **Tables S1-S4** in the Supplementary Appendix).^12–16^ In each simulation, we calculated the sum of square errors between the model output and data and selected the top 10% with the least square errors to generate 95% confidence intervals. The detailed calibration procedure is described in the Supplementary Appendix. All analyses and simulations were performed in MATLAB R 2019b.

**Figure 2.**
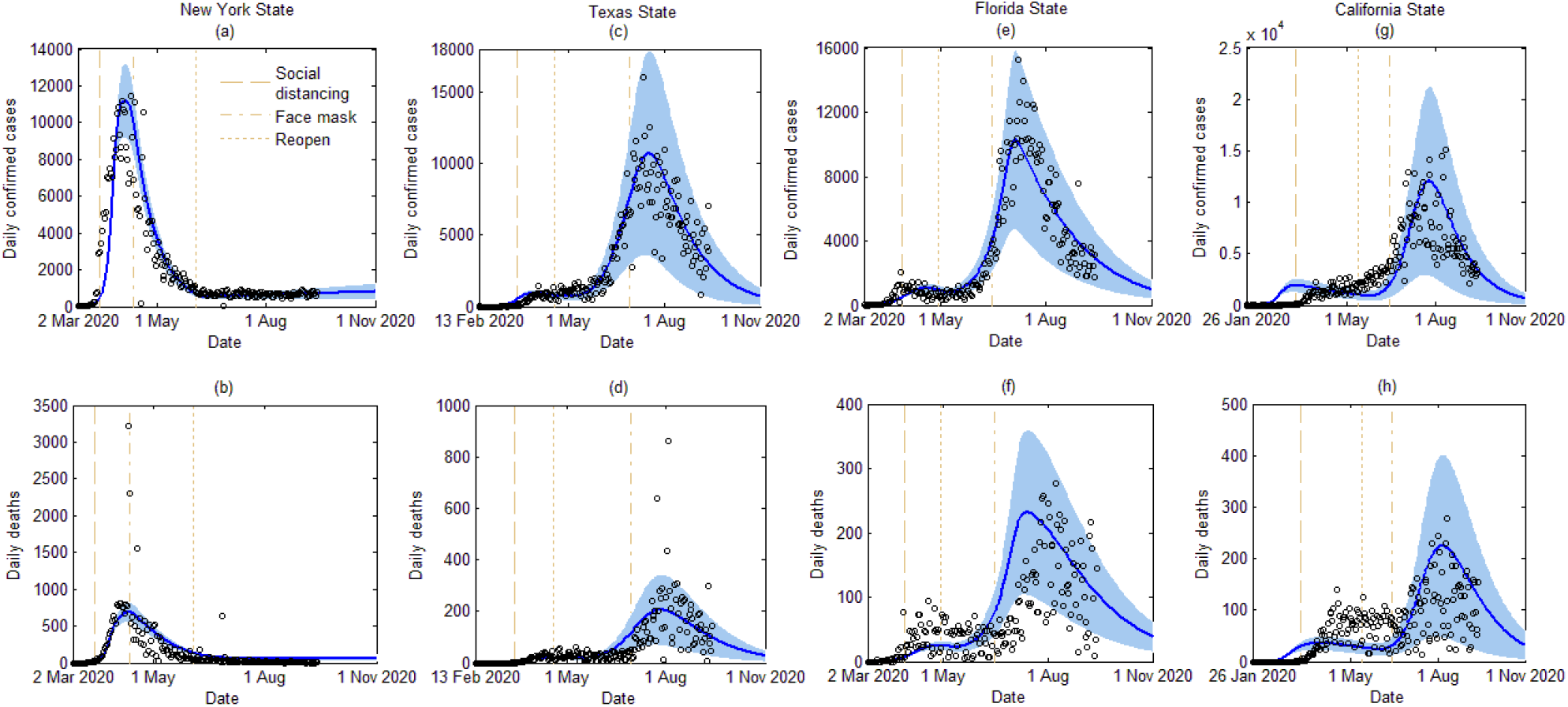
Model calibration and data fitting based on reported confirmed COVID-19 cases and deaths in four states, i.e., New York (a-b), Texas (c-d), Florida (e-f), and California (g-h). The blue areas denote 95% confidence intervals. Dashed lines, dash-dot lines, and dotted lines denote the social distancing order (public person-to-person contact rates decreased), face mask order, and reopening (public person-to-person contact rates recovered to no more than 100% of the pre-pandemic level) policies that were implemented in each state, respectively.

### Construction of scenarios

We projected the number of cases and deaths under the following four scenarios: (1) the no vaccine scenario in which social distancing restrictions are relaxed (public person-to-person contact rates recovered to 100% of the pre-pandemic level), but the vaccine has not been developed; (2) the vaccine and face mask scenario in which people are vaccinated (with different effectiveness and coverage) while social distancing restrictions are relaxed, and the baseline face mask use rate is maintained; (3) the vaccine and reduced face mask scenario in which people are vaccinated while social distancing restrictions are relaxed, and half of the baseline face mask use rate is maintained; (4) the vaccine and no face mask scenario in which people are vaccinated while social distancing restrictions are relaxed and face masks are no longer used.

We assume that natural and vaccine-induced immunity would last for at least one year. We then calculated the number of averted infections and deaths after one year for scenarios (2)-(4), compared to scenario (1), and plotted them as a function of vaccine effectiveness and coverage. The threshold of vaccination curve was defined as the combination of vaccine effectiveness and coverage such that social distancing restrictions may be relaxed while the COVID-19 epidemic can be retained at a sustainably low endemic level or eliminated.

## RESULTS

Our results (**Figures 3-4**) show that, in the absence of a vaccine, if social distancing restrictions were relaxed while the current face mask use rate was maintained, there would be 0.8-4 million new infections and 15,000-240,000 new deaths across the four states within one year. If the current face mask use rate was maintained, introducing an effective vaccine would always decrease the number of infections and deaths. Greater vaccine effectiveness and/or coverage rate would lead to more averted infections and deaths. However, if the face mask use rate decreased by 50%, a low vaccine effectiveness and coverage rate may not be enough to eliminate or suppress the pandemic to a low level without further major outbreaks. If people no longer wore face masks, a greater vaccine effectiveness and coverage rate would be needed to suppress the pandemic. We define vaccine effectivenesses of 50%, 80%, and 100% as weak, moderate, and strong, respectively, and present state-specific results next.

**Figure 3.**
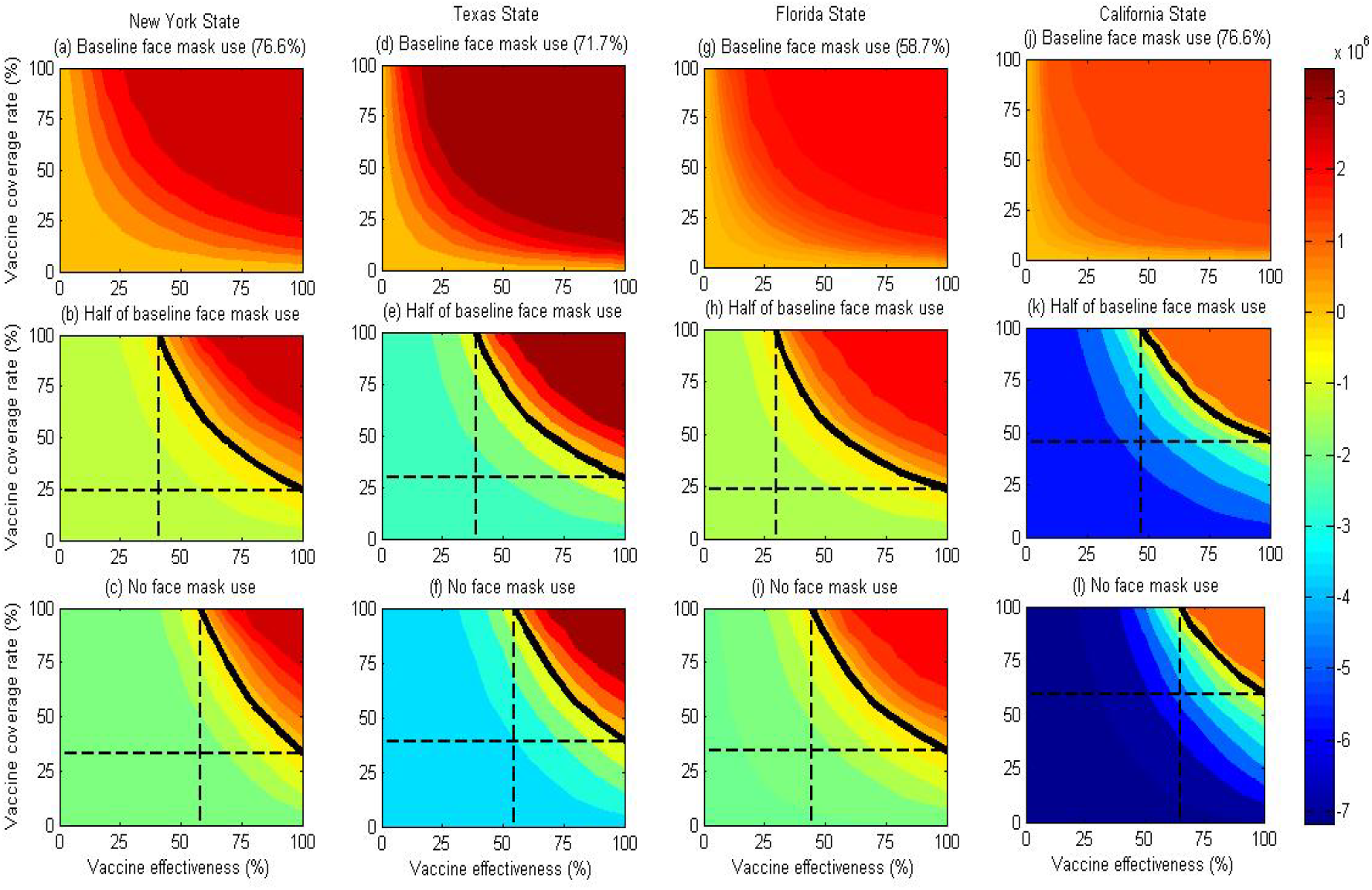
Contour plots of averted infections as a function of vaccine effectiveness and vaccine coverage rate in four states when social distancing is relased to pre-pandemic level shortly after the commencement of vaccination. The first row is maintaining face mask use at baseline levels, the second row to reducing face masks use to half of the baseline level and the third row is no face mask use. The black solid isoclines indicate the threshold that the number of averted infections is zero. The black dashed lines correspond to the minimal vaccine effectiveness and vaccine coverage rate when the number of averted infections is zero.

**Figure 4.**
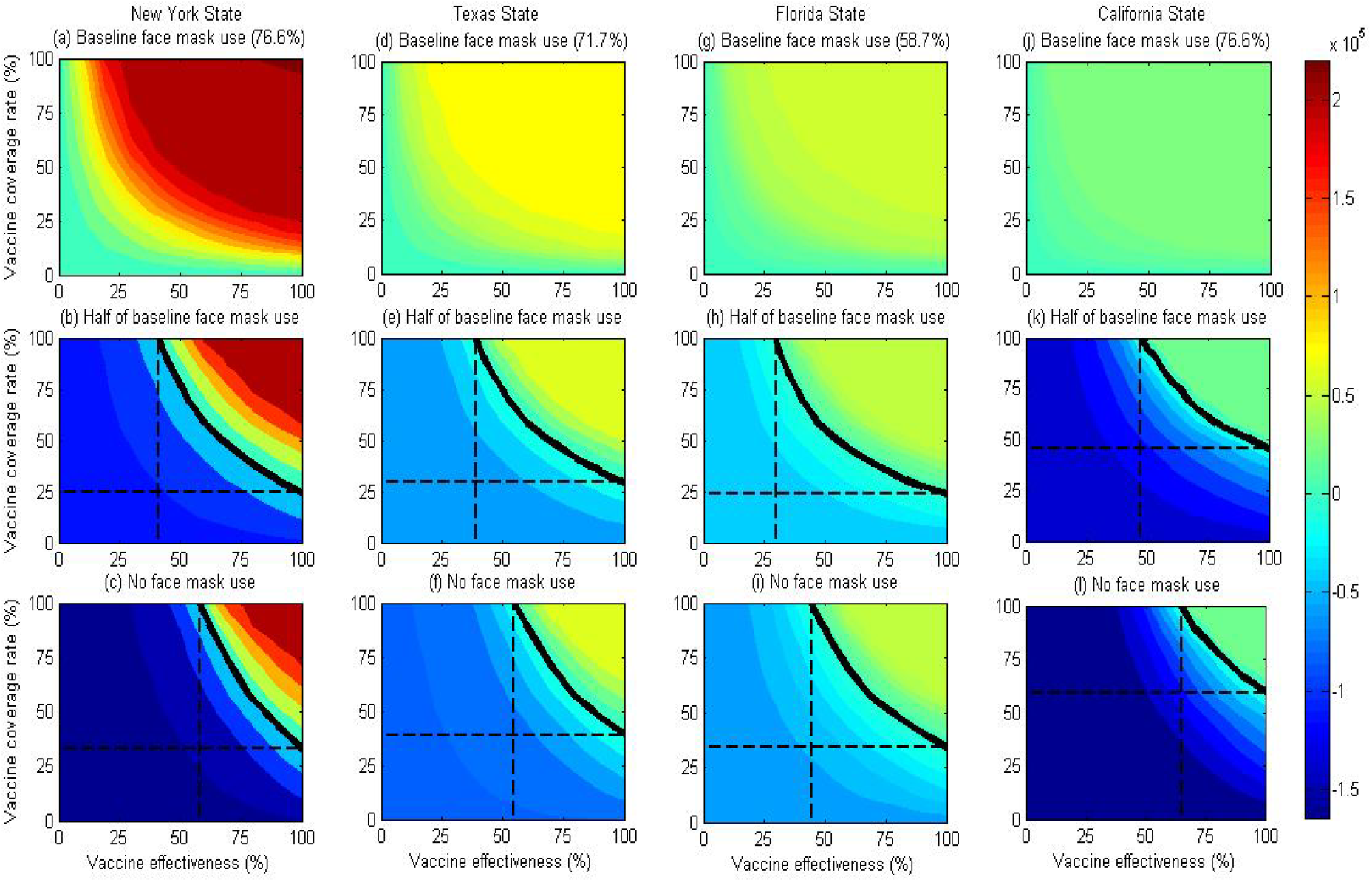
Contour plots of averted deaths as a function of vaccine effectiveness and vaccine coverage rate in four states when relaxing social distancing to pre-pandemic level and maintaining face mask use to the baseline level (the first line), half of the baseline level (the second line), and no use (the third line), compared with no vaccine. The black solid isoclines indicate the threshold that the number of averted deaths is zero. The black dashed lines correspond to the minimal vaccine effectiveness and vaccine coverage rate when the number of averted deaths is zero.

### New York

Figures 3a and 4a show that, in the state of New York, if the current face mask use was maintained and vaccine effectiveness was weak (50% effectiveness), 2.49 (95% CI, 2.37-2.61) million infections and 203,445 (189,366-217,525) deaths could be averted under 50% vaccine coverage, 2.65 (2.51-2.79) million infections and 216,290 (197,944-234,635) deaths could be averted under 75% vaccine coverage, and 2.68 (2.53-2.83) million infections and 218,854 (199,337-238,372) deaths could be averted under 100% coverage. Increasing the vaccine effectiveness would avert more infections and deaths. For example, a moderate vaccine (80% effectiveness) could avert 2.66 (2.51-2.80) million infections and 216,698 (198,143-235,252) deaths under 50% coverage and a strong vaccine (100% effectiveness) could avert 2.67 (2.52-2.82) million infections and 218,150 (198,929-237,372) deaths under the same coverage.

Figures 3b and 4b showed that decreasing face mask use to 50% of the current use would require greater vaccine effectiveness and coverage to suppress the pandemic. The threshold of vaccination curve showed that if the vaccine effectiveness is weak or moderate, the coverage should be greater than 77.1% or 38.0%, respectively, to suppress the pandemic. Deferring the vaccine rollout by two months would require coverage of 75.3% and 37.7% under a weak or moderate vaccine, respectively, to suppress the pandemic (Figures S4-S5 in the Supplementary Appendix).

Figures 3c and 4c showed that if people did not wear face masks and the vaccine effectiveness was weak, even 100% vaccine coverage would not suppress the pandemic. If the vaccine effectiveness was moderate or strong, the vaccine coverage should be greater than 55.4% or 33.2% to suppress the pandemic. Deferring the rollout of vaccine by two months would require 54.8% and 33.2% coverage to suppress the pandemic under moderate or strong effectiveness. (Figures S4-S5 in the Supplementary Appendix).

### Texas

In the state of Texas, if the current face mask use rate was maintained and the vaccine had a weak effectiveness, 50% vaccine coverage could avert 3.36 (2.87-3.85) million infections and 72,732 (61,309-84,156) deaths, 75% coverage could avert 3.41 (2.91-3.91) million infections and 73,719 (62,088-85,349) deaths, and 100% coverage could avert 3.42 (2.92-3.92) million infections and 73,984 (62,340-85,628) deaths (Figures 3d and 4d). A moderate vaccine would avert avert 3.41 (2.91-3.91) million, 3.42 (2.92-3.92) million, 3.43 (2.93-3.92) million infections, and 73,754 (62,121-85,387), 73,999 (62,352-85,646), 74,127 (62,484-85,770) deaths, respectively, under 50%, 75%, 100% vaccine coverage.

If the face mask use decreased by 50% (Figures 3e and 4e) or there was no face mask use (Figures 3f and 4f), greater vaccine effectiveness and coverage would be needed to suppress the pandemic. For example, if the vaccine effectiveness was weak, the vaccine coverage should be greater than 74.6% to suppress the pandemic under 50% face mask use. Deferring the rollout of vaccine by two months would require a 67.9% coverage to suppress the pandemic (Figures S4-S5 in the Supplementary Appendix). When no face mask was used, and the vaccine was weak, even 100% coverage would not suppress the pandemic in Texas.

### Florida

In the state of Florida, if the current face mask use rate was maintained and the vaccine was weak, 50% coverage could avert 2.01 (1.54-2.48) million infections and 55,001 (41,197-68,805) deaths, 75% coverage could avert 2.04 (1.58-2.51) million infections and 56,034 (42,216-69,852) deaths, and 100% coverage could avert 2.06 (1.59-2.52) million infections and 56,406 (42,661-70,152) deaths (Figures 3g and 4g). If the vaccine was moderate, then 50%, 75%, 100% vaccine coverage rates could avert 2.05 (1.58-2.51) million, 2.06 (1.59-2.52) million, 2.06 (1.60-2.53) million infections, and 56,065 (42,247-69,882), 56,402 (42,652-70,153), 56,610 (42,918-70,301) deaths, respectively. If the vaccine was strong, then 50%, 75%, 100% coverage could avert 2.05 (1.59-2.52) million, 2.06 (1.60-2.52) million, 2.07 (1.61-2.53) million infections, and 56,260 (42,476-70,044), 56,496 (42,770-70,221), 56,668 (42,995-70,342) deaths, respectively.

If the face mask use decreased by 50% and the vaccine effectiveness was weak (Figures 3h and 4h), the threshold of vaccination curve showed that the coverage should be greater than 55.0% to suppress the pandemic. If the vaccine effectiveness was moderate or strong, the vaccine coverage should be greater than 32.2% and 23.4%, respectively, to suppress the pandemic. Deferring the rollout of vaccine by two months with 50% of the current face mask use would require 30.8% and 23.0%, respectively, to suppress the pandemic (Figures S4-S5 in the Supplementary Appendix). If no face mask was used, the required vaccine coverage rates would be 87.8% and 47.8% under the moderate and strong effectiveness, respectively, to suppress the pandemic (Figures 3i and 4i).

### California

In the state of California, if the current face mask use rate was maintained and the vaccine was weak, 50% coverage could avert 1.42 (0.761-2.088) million infections and 28,757 (14,789-42,726) deaths, 75% coverage could avert 1.44 (0.77-2.11) million infections and 29,016 (14,896-43,136) deaths, and 100% coverage could avert 1.44 (0.77-2.11) million infections and 29,146 (14,959-43,332) deaths (Figures 3j and 4j). If the vaccine was moderate, then 50%, 75%, 100% coverage could avert 1.44 (0.77-2.11) million, 1.44 (0.77-2.11) million, 1.45 (0.77-2.12) million infections, and 29,024 (14,900-43,149), 29,143 (14,957-43,330), 29,228 (15,000-43,456) deaths, respectively. A strong vaccine would lead to 1.44 (0.77-2.11) million averted infections and 29,091 (14,930-43,252) averted deaths under 50% coverage, 1.44 (0.770-2.12) million averted infections and 29,181 (14,975-43,386) averted deaths under 75% coverage, and 1.45 (0.77-2.12) million averted infections and 29,254 (15,014-43,494) averted deaths under 100% coverage.

If the face mask use decreased by 50%, and the vaccine was weak, the vaccine coverage should be greater than 94.2% to suppress the pandemic (Figures 3k and 4k). If the vaccine was moderate or strong, the vaccine coverage should be greater than 56.8% and 45.7%, respectively, to suppress the pandemic. If no face mask was used, and the vaccine effectiveness was weak, even 100% coverage would not decrease the number of infections or deaths (Figures 3l and 4l). If the vaccine effectiveness was moderate, the vaccine coverage of great than 77.8% would be required to suppress the pandemic based on the threshold of vaccination curve. If the vaccine effectiveness was strong, less vaccine coverage (58.0%) would be required. Similar to the other states, deferring the rollout of vaccine would only moderately decrease the vaccine coverage required to suppress the pandemic in California.

## DISCUSSION

Our study addressed important questions related to what would be needed to suppress COVID-19 in New York, Texas, Florida, and California under different scenarios of vaccine effectiveness, uptake, and face mask use. The results suggest that the number of COVID-19 cases would decrease in the four most severely affected states in the US under the current approach of relying primarily on social distancing and mask use. However, if these measures are relaxed before an effective vaccine becomes available, the epidemic will likely rebound with further major outbreaks.^17^ So far, all four states have partially or fully reopened their economies, but face mask use in public space remains mandatory or recommended. Our study suggests that if face mask use was maintained at the current level, vaccination if it were only moderately effective would result in lower numbers of new infections and deaths even when social distancing returned to normal. If face mask usage was halved in these states, then a weak vaccine (50% effectiveness) would require 55-94% population coverage to suppress the epidemic, whereas a moderate vaccine (80% effectiveness) would require 32-57% population coverage and a strong vaccine (100% effectiveness) would require only 24-46% population coverage to suppress COVID-19. In all scenarios social distancing was assumed to return to pre-epidemic levels. In contrast, if face mask usage is reduced to zero then a weak vaccine would not provide substantial protection, and further outbreaks are anticipated, but a moderate vaccine may suppress the epidemic with vaccination coverage of 48-78%, and a strong vaccine would require 33-58% coverage.

For social distancing to be returned to the pre-pandemic level in the four most severely COVID-19 affected states in the US, at least half of its population needs to receive a vaccine with moderate effectiveness. However, the state of California, in particular, will need a vaccine coverage of close to 80% to suppress the COVID-19 epidemic, such that both social distancing restrictions and the requirement for face mask use can be relaxed. The requirement of higher vaccination coverage in California is likely due to a higher proportion of susceptible individuals compared to the other three states. In other words, the prevalence of infected individuals with natural immunity is lower in California.

A recent study indicated that vaccinating 82% of the US population with a vaccine of 80% effectiveness is necessary to achieve herd immunity and eliminate COVID-19.^18^ A separate study also also confirmed this estimate and demonstrated that with a moderately effective vaccine (80% effectiveness), the threshold coverage for the US population would be at least 60-80% when the virus reproduction number varied between 2.5-3.5.^19^ Given that the willingness to take a COVID-19 vaccine in the US has been estimated at only 58%,^20,21^ only a strong vaccine with high effectiveness of nearly 100% would be sufficient to suppress the epidemic alone and enable relaxation of social distancing and face mask requirement. If a strong vaccine is not attainable, a moderate vaccine and maintaining a relatively low face mask usage of 30-40% would also be a plausible alternative to achieve the same target. That is, vaccination combined with a modest level of non-pharmaceutical measures, such as face mask use in common public spaces (shopping malls and transportation), may be a viable option to continue suppressing the epidemic in the long term.

Our findings demonstrate that the timing of a vaccination rollout may only have a small impact on the threshold vaccination coverage. Deferring vaccination rollout by one or two months did not substantially change the required threshold coverage. However, if a safe and effective vaccine becomes available, it should be delivered to the population as early as possible to support the economic recovery of the country.^22–24^ Despite reopening the economy in these states, the restrictions related to interstate and international travel have significantly disrupted the recovery of the US economy.^25^ Only an effective vaccination program is able to counteract these restrictions.^26–28^

Our study has a number of limitations. First, our model did not take into consideration the age structure of the population because data are currently not available on the different effects of a potential vaccine across age groups. Second, the model did not distinguish between vaccine types (e.g., inactivated, live attenuated, recombinant protein) and the doses of vaccine. We used the average vaccine effectiveness to address the difference of vaccine types and doses in the model. Third, the model did not consider the lag time required for the vaccine to become effective and assumed an immediate immune response and protection after vaccination. Our sensisivity analyses showed that one- or two-month delay of immune response would have little impact on the results. Fourth, we assumed that the vaccine protection lasts for at least one year and, thus, did not project the epidemic beyond one year. If the vaccine protection duration was shorter than one year, it would need larger vaccine coverage to suppress the epidemic. Finally, the model did not consider issues related to vaccine availability, distribution, and the cost-effectiveness of vaccination,^22,24,29^ which would be important future research directions when more data (e.g., vaccine cost, quality of life for COVID-19 patients) become available.

## CONCLUSIONS

Our study indicates that for people to return to their pre-pandemic normal life, a potential vaccine needs to have moderate effectiveness of 80% and cover at least 50-80% for the four most severely affected states in the US. Maintaining a 30-40% face mask use would reduce the required vaccine coverage below the willingness level of vaccination of the US population. Delaying vaccination rollout for 1-2 months would not substantially alter the epidemic trend if the current non-pharmaceutical interventions were maintained. The findings from this study can inform the planned rollout of COVID-19 vaccines and the continued implementation of non-pharmaceutical interventions such as social distancing and mask use mandates.

## Data Availability

Data are publicly available.

## References

1. Miller IF, Becker AD, Grenfell BT, Metcalf CJE. Disease and healthcare burden of COVID-19 in the United States. Nat Med. 2020;26(8):1212–1217.

2. Chen JT, Krieger N. Revealing the Unequal Burden of COVID-19 by Income, Race/Ethnicity, and Household Crowding: US County Versus Zip Code Analyses. J Public Health Manag Pract. Published online 2020.

3. Weinberger DM, Chen J, Cohen T, et al. Estimation of excess deaths associated with the COVID-19 pandemic in the United States, March to May 2020. JAMA Intern Med. 2020;180(10):1336–1344.

4. Bartsch SM, Ferguson MC, McKinnell JA, et al. The Potential Health Care Costs And Resource Use Associated With COVID-19 In The United States: A simulation estimate of the direct medical costs and health care resource use associated with COVID-19 infections in the United States. Health Aff (Millwood). Published online 2020:10–1377.

5. JHU. Johns Hopkins University Coronavirus Resource Center.; 2020. https://coronavirus.jhu.edu/

6. Poland GA, Ovsyannikova IG, Kennedy RB. SARS-CoV-2 immunity: review and applications to phase 3 vaccine candidates. The Lancet. Published online 2020.

7. Jackson LA, Anderson EJ, Rouphael NG, et al. An mRNA vaccine against SARS-CoV-2—preliminary report. N Engl J Med. Published online 2020.

8. Zhu F-C, Guan X-H, Li Y-H, et al. Immunogenicity and safety of a recombinant adenovirus type-5-vectored COVID-19 vaccine in healthy adults aged 18 years or older: a randomised, double-blind, placebo-controlled, phase 2 trial. The Lancet. 2020;396(10249):479–488.

9. Lyu W, Wehby GL. Community Use Of Face Masks And COVID-19: Evidence From A Natural Experiment Of State Mandates In The US: Study examines impact on COVID-19 growth rates associated with state government mandates requiring face mask use in public. Health Aff (Millwood). 2020;39(8):1419–1425.

10. Chu DK, Akl EA, Duda S, et al. Physical distancing, face masks, and eye protection to prevent person-to-person transmission of SARS-CoV-2 and COVID-19: a systematic review and meta-analysis. The Lancet. Published online 2020.

11. The New York Times and Dynata. Mask-Wearing Survey Data. New York Times; 2020. https://github.com/nytimes/covid-19-data/blob/master/mask-use/mask-use-by-county.csv

12. Zhang J, Litvinova M, Liang Y, et al. Changes in contact patterns shape the dynamics of the COVID-19 outbreak in China. Science. Published online 2020.

13. Jarvis CI, Van Zandvoort K, Gimma A, et al. Quantifying the impact of physical distance measures on the transmission of COVID-19 in the UK. BMC Med. 2020;18:1–10.

14. Zhang L, Shen M, Ma X, et al. What Is Required to Prevent a Second Major Outbreak of SARS-CoV-2 upon Lifting Quarantine in Wuhan City, China. The Innovation. 2020;1(1):100006.

15. Shen M, Peng Z, Guo Y, et al. Assessing the effects of metropolitan-wide quarantine on the spread of COVID-19 in public space and households. Int J Infect Dis. Published online 2020.

16. Shen M, Peng Z, Xiao Y, Zhang L. Modelling the epidemic trend of the 2019 novel coronavirus outbreak in China. Innovation. 2020;3. doi:https://doi.org/10.1016/j.xinn.2020.100048

17. Zhang L, Tao Y, Shen M, Fairley CK, Guo Y. Can self-imposed prevention measures mitigate the COVID-19 epidemic? PLoS Med. 2020;17(7):e1003240.

18. Gumel AB, Iboi EA, Ngonghala CN. Will an imperfect vaccine curtail the COVID-19 pandemic in the US? medRxiv. Published online 2020.

19. Bartsch SM, O’Shea KJ, Ferguson MC, et al. Vaccine efficacy needed for a COVID-19 coronavirus vaccine to prevent or stop an epidemic as the sole intervention. Am J Prev Med. 2020;59(4):493–503.

20. Reiter PL, Pennell ML, Katz ML. Acceptability of a COVID-19 vaccine among adults in the United States: How many people would get vaccinated? Vaccine. 2020;38(42):6500–6507.

21. Fisher KA, Bloomstone SJ, Walder J, Crawford S, Fouayzi H, Mazor KM. Attitudes Toward a Potential SARS-CoV-2 Vaccine: A Survey of US Adults. Ann Intern Med. Published online 2020.

22. Bollyky TJ, Gostin LO, Hamburg MA. The equitable distribution of COVID-19 therapeutics and vaccines. JAMA. Published online 2020.

23. Adalja AA, Toner E, Inglesby TV. Priorities for the US health community responding to COVID-19. Jama. 2020;323(14):1343–1344.

24. Persad G, Peek ME, Emanuel EJ. Fairly prioritizing groups for access to COVID-19 vaccines. JAMA. Published online 2020.

25. Studdert DM, Hall MA, Mello MM. Partitioning the Curve—Interstate Travel Restrictions During the Covid-19 Pandemic. N Engl J Med. Published online 2020.

26. DeRoo SS, Pudalov NJ, Fu LY. Planning for a COVID-19 Vaccination Program. JAMA. Published online 2020.

27. Corey L, Mascola JR, Fauci AS, Collins FS. A strategic approach to COVID-19 vaccine R&D. Science. 2020;368(6494):948–950.

28. Fauci AS, Lane HC, Redfield RR. Covid-19—Navigating the Uncharted. Mass Medical Soc; 2020.

29. Gayle H, Foege W, Brown L, Kahn B. Framework for equitable allocation of COVID-19 vaccine. Natl Acad Med. Published online 2020. https://www.arfda.com/Portals/0/Content/News/Files2020/NASFrameworkCOVIDVaccine.pdf

