## supplementary appendix for "Projected COVID-19 epidemic in the United States in the context of the effectiveness of a potential vaccine and implications for social distancing and face mask use"

This supplementary document describes in detail the mathematical model and estimation of parameters presented in the main text.

**1. Model formulation**

We proposed a dynamic compartmental model to describe the transmission of COVID-19 in four states (New York, Texas, California, Florida) in United States. The population in each state is divided into ten compartments (**Figure S1**): susceptible individuals (S), vaccinated individuals (V), latent infections (E), asymptomatic infections (A), undiagnosed infections with mild/moderate (I_1_) and severe/critical symptoms (I_2_), diagnosed infections with mild/moderate (T_1_) and severe/critical symptoms (T_2_), recovered (R) and deceased (D) cases. The total population size is denoted by N, where N=S+V+E+A+I_1_+I_2_+T_1_+T_2_+R.

S

E

I_1_

T_1_

R

D

I_2_

T_2_

A

$$\Lambda$$

$$k_{1}\rho$$

$$k_{1}(1-\rho)$$

$$k_{2}$$

$$k_{3}$$

$$\gamma_{0}$$

$$\gamma_{0}$$

$$\alpha_{1}$$

$$\alpha_{2}$$

$$\gamma_{1}$$

$$\gamma_{2}$$

$$\mu_{1}$$

$$\mu_{2}$$

V

$$\Lambda_{V}$$

$$w$$

**Figure S1**. A schematic flow diagram of the transmission of COVID-19.

**Model without vaccine**

We first construct the model in the absence of vaccine and then extend it by including the vaccinated class. Susceptible individuals become infected by contacts with latent, asymptomatic and undiagnosed infectious individuals with symptoms in the public settings (e.g. public transportations, supermarkets, offices, etc) and household (home or other private settings). The total force of infection $\Lambda$ is given by the sum of forces of infections via these routes. That is,

$\Lambda= \Lambda_{pub}+\Lambda_{pri}.$	⑴

For each of route of transmission,

(a) public contacts with susceptible

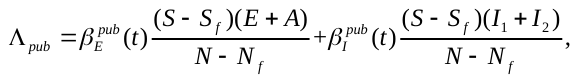
 ⑵

(b) household contacts with susceptible

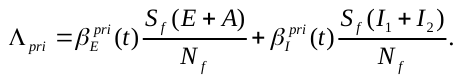
 ⑶

where

$\beta_{I}^{pub}(t)=\beta m_{1}(t)(1-\theta_{1}p_{1}(t))(1-\theta_{2}q(t)),\beta_{E}^{pub}(t)=(1-\varepsilon)\beta_{I}^{pub}(t),$	$\beta_{I}^{pri}\left( t \right)=\beta m_{2}\left( t \right)\left( 1-\theta_{1}p_{2} \right)(1-\theta_{2}q(t)),\beta_{E}^{pri}\left( t \right)=\left( 1-\varepsilon\right)\beta_{I}^{pri}\left( t \right).$	⑷

Here $\beta$ denotes the probability of transmission per contact with the infectious individuals with symptoms. We assumed that for contacts with the latent and asymptomatic individuals this probability is lower, i.e. $(1-\varepsilon)\beta$ where $0\leq\varepsilon\leq1$ denotes the reduction in per-act transmission probability. The parameters *m*_1_(*t*) and *m*_2_(*t*) represent the average number of daily person-to-person contacts in the public settings and household, *p*_1_(*t*) and *p*_2_ denote the proportion of mask usage in the public settings and household, respectively, and $\theta_{1}$ is the effectiveness of face mask/respirators in infection prevention. Let $q(t)$ and $\theta_{2}$ denote the proportion of handwashing (assumed the same in public settings and household) and the effectiveness of handwashing in infection prevention.

*Estimation of the population size and number of susceptibles for each route of transmission*

For household contacts, the overall population size (*N_f_*) is estimated as the total number of households members that are at risk of COVID-19 infection, whereas the number of susceptible households members (*S_f_*) is the difference between *N_f_* and the number of infected individuals in these households. We assumed that the number of the households at risk of infection is the same as the number of individuals infected in public settings because the probability of two or more household members being infected at the same time but at different public venues is very small. Hence, the entry of *N_f_* is$r\Lambda_{pub}$, where *r* is the average number of household members in a US family and $\Lambda_{pub}$ is the number of individuals infected in public settings as shown in eq. (2). We assumed that the infected family members become recovered after the mean period $1/\xi$. Further, the entry of susceptible household members *S_f_* is $\left( r-1 \right)\Lambda_{pub}-\Lambda_{pri}$, where $\Lambda_{pri}$ denotes the number of infected household members through household transmission as shown in eq. (3).

For public contacts, the overall population size is the number of residents (*N*) in the state minus the overall population size (*N_f_*) in household contacts, whereas the number of susceptibles is the number of individuals free of COVID-19 infection (*S*) minus the susceptibles (*S_f_*) in household contacts.

*Modeling disease progression*

Individuals in the incubation period (*E*) progress to the infectious compartment with mild/moderate symptoms or asymptomatic compartment at a rate *k*_1_ and the probability that an individual is asymptomatic is $\rho$. Infectious individuals with mild/moderate and severe/critical symptoms are diagnosed and treated at the rates $\alpha_{1}$and $\alpha_{2}$, respectively. We assume these diagnosed individuals are isolated strictly and could not further infect others. Undiagnosed and diagnosed mild/moderate cases progress to the severe/critical stage at the rates $k_{2}$and $k_{3}$, respectively. Asymptomatic infections and undiagnosed mild/moderate cases are assumed to recover naturally at the rate $\gamma_{0}$. Diagnosed mild/moderate and severe/critical cases will recover at the rates $\gamma_{1}$ and $\gamma_{2}$, respectively. Undiagnosed and diagnosed severe/critical cases will die due to the disease at the rates $\mu_{1}$ and $\mu_{2}$, respectively. The model is described by the following system of ordinary differential equations:
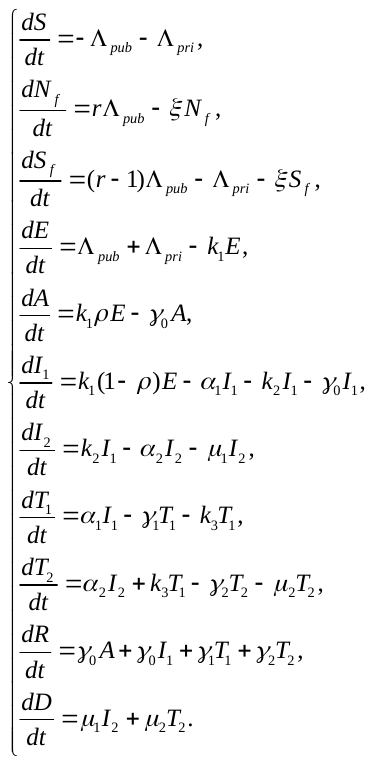
⑸

The cumulative number of deaths is tracked by the last equation of *D* in eq. (5) and the cumulative number of confirmed cases *C* is governed by the equation

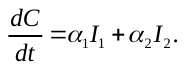
 ⑹

**Model with vaccine**

In this section, we assume that the susceptible would be vaccinated by a COVID-19 vaccine. The vaccination rate is *w* and the effectiveness of the vaccine is $\varepsilon_{V}$. Susceptible and vaccinated individuals become infected by contacts with latent, asymptomatic and undiagnosed infectious individuals with symptoms in the public settings (e.g. public transportations, supermarkets, offices, etc) and household (home or other private settings). The total force of infection $\Lambda$ and $\Lambda_{V}$ for susceptible and vaccinated individuals are given by the sum of forces of infections via these routes. That is,

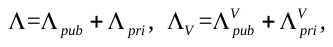
 (7)

(a) public contacts with susceptible

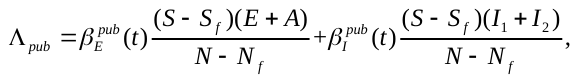
 (8)

(b) household contacts with susceptible

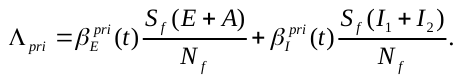
 (9)

(c) public contacts with vaccinated individuals

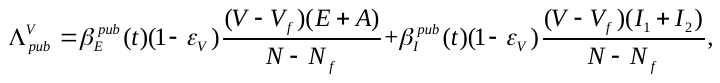
 (10)

(d) household contacts with vaccinated individuals

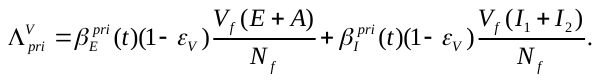
 (11)

where *N_f_* is the total number of households members that are at risk of COVID-19 infection, *S_f_* is the number of susceptible households members, and *V_f_* is the number of vaccinated households members.

This, we obtain the full model in the presence of vaccine as follow:

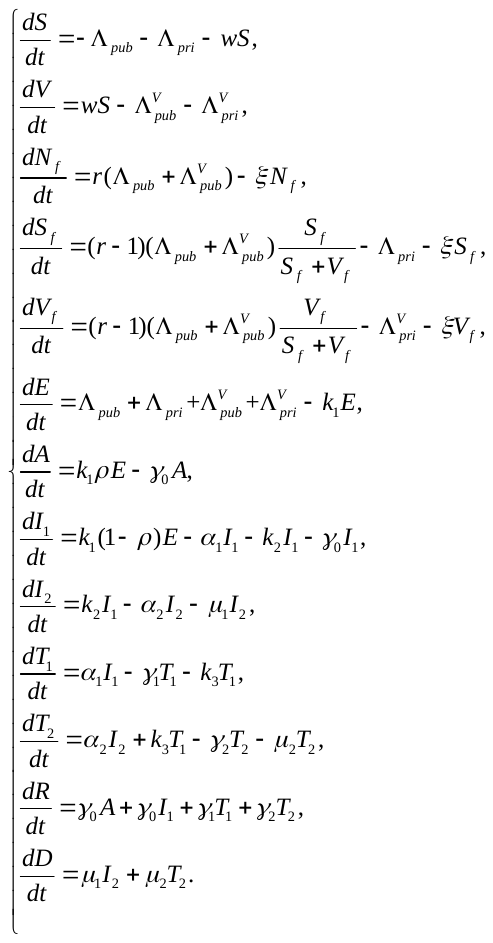
(12)

**2. Data and parameter estimation**

We collected the data on the number of daily and cumulative confirmed cases and deaths of four states, i.e., New York, Texas, Florida, and California, from January 26 to September 15, 2020 from the Johns Hopkins University Coronavirus resource center.^1^ The mean incubation time for COVID-19 is about 5.2 days (1/*k*_1_=5.2).^2^ The mean time from the onset of symptoms to severe/critical symptoms was 10 days (1/*k*_2_=10).^3^ The mean number of members in a US household is 3.14, so we assumed *r*=4.^4^ The average period from the onset of symptoms to diagnosis is 7 days and from the onset of symptoms to recovery for those with mild/moderate symptoms is 2 weeks in China,^5^ which indicates that the average recovery period for diagnosed mild/moderate cases is 14-7=7 days, so we assume the same average recovery period in US, i.e., 1$/\gamma_{1}$=7. Similarly, the average period from the onset of symptoms to recovery for those with severe/critical symptoms is 3 weeks,^6^ we have $\frac{1}{\gamma_{2}}+\frac{1}{\alpha_{2}}=21$. We assumed that the duration of recovery for infected family members is 4 weeks ($1/{\xi=28)}$.^7^ The probability of transmission per contact with latent and asymptomatic individuals is assumed to be 25% ($1-\varepsilon$=0.25) of that with infectious symptomatic individuals.^8^

We describe the detailed parameter estimation process for New York State as an example and it is similar for other three states. The data from China^9^ and UK^10^ show that the number of daily contacts decreased by 80% and 74%, respectively after the city quarantine. Similarly, we assumed that the average number of daily contacts in the public settings $m_{1}(t)$ can be reduced by up to 80% in New York State in the base case,^11^ described by a decreasing logistic function${m_{1}\left( t \right)=m}_{ini}+\frac{0.2m_{ini}-m_{ini}}{1+exp(-m_{0}(t-t_{ini} ))}$, where $m_{ini}$ is the background daily contact number in the public settings before the state was put “on pause”,^12^ $m_{0}$ is the change rate of contact number, $t_{ini}$ is the time when contact number is the half of initial and minimal contact number in the public settings and these three parameters will be estimated by model fitting. Home confinement led to double contact rate than the pre-quarantine level.^13^ We assumed that the average number of daily contacts in a household increased from 4 to 8, described by an increasing logistic function $m_{2}\left( t \right)=4+\frac{8-4}{1+exp(-m_{0}(t-t_{ini}))}$. After reopening since June 7, 2020, the contact number in the public settings increased and we assumed the contact in households decreased simultaneously, i.e., ${m_{1}\left( t \right)=0.2m}_{ini}+\frac{m_{1,reopen}\times0.2m_{ini}-{0.2m}_{ini}}{1+\exp(-m_{0}(t-t_{reopen}))}$ and $m_{2}\left( t \right)=8+\frac{m_{2,reopen}\times8-8}{1+\exp(-m_{0}(t-t_{reopen}))}$, where two parameters $m_{1,reopen}, m_{2,reopen}$ will be estimated by model fitting.

The proportion of handwashing in the US was assumed to increase during the outbreak.^14^ We assumed a logistic growth for this percentage (**Figure S2b**), i.e. ${q\left( t \right)=q}_{ini}+\frac{\bar{q}-q_{ini}}{1+exp(-m_{0}(t-t_{ini}))}$ where $q_{ini}$ is the base proportion of handwashing before the outbreak, and $\bar{q}$ is the maximum proportion of handwashing during the outbreak. A survey on handwashing culture from 63 countries showed that about 77% of people in US had a habit of automatic handwashing in routine life,^14^ so we chose $p_{ini}=77\%$ as the base value of the proportion of handwashing in the absence of COVID-19. Another online survey in US showed that 88.27-99% of people washed hands during the epidemic,^15^ so we assumed $\bar{q}=95\%$ as the maximum proportion of handwashing in New York State. The effectiveness of handwashing in preventing infection ($\theta_{2}$) is chosen to be 42% (50-95%), based on a review against respiratory infections.^16^

US CDC recommended face mask use in April 3, and after two weeks New York State implemented the Executive Order on face mask use that required all residents over age 2 must wear masks or face coverings when they're in public and social distancing isn't possible.^17^ We assumed its use coverage increased with a logistic growth, i.e. ${p_{1}\left( t \right)=p}_{ini}+\frac{\bar{p}-p_{ini}}{1+exp(-(t-t_{p}))}$ where $p_{ini}=0$ is the base proportion of face mask usage in the public settings before CDC recommendation, $\bar{p}=76.6\%$ is the adjusted state-level maximum proportion of face mask usage in the public settings after the Executive Order on face mask use based on county-level data of always wearing a mask^18^ and demographic data in each county,^19^ and $t_{p}=42$ is the time when the proportion of face mask usage is the half of base and maximum proportion. The coverage ratio of face mask use in private settings $p_{2}$ is set to zero.^20^ The effectiveness of face mask in preventing infection ($\theta_{1}$) is chosen to be 85% (66-93%), based on a meta-analysis against COVID-19.^21^ We varied the effectiveness of face mask from 0 to 100% in sensitivity analysis.

The total population size in New York State is 19,542,209.^19^ The initial values of the disease states are given by T_1_(0)=0, T_2_(0)=0, R(0)=0, D(0)=0, N(0)= 19,542,209, $N_{f}\left( 0 \right)=rC\left( 0 \right)=4$, and $S_{f}\left( 0 \right)=(r-1)C\left( 0 \right)=3$. We left E(0),A(0), I_1_(0), I_2_(0) to be estimated by the fitting.

The total population size in Texas, Florida, and California States are 28,701,845, 21,299,325, and 39,557,045,^19^ respectively, and the initial values of the disease states can be obtained similarly. The parameter estimation process for these three states are also similar to that for New York State (see **Table S2-S4**).

We calibrated the model by the daily and cumulative confirmed cases and deaths from 26^th^ January to 15^th^ September, 2020 for the four states by using a nonlinear least-squares method (**Figure 2**). The unknown parameters (**Table S1-S4**) were sampled within their ranges by the Latin hypercube sampling method and repeated 1000 times. For every simulation, we calculated the sum of square errors between the model output and data, and selected the top 10% with the least square errors to generate 95% confidence intervals.

As an illustrative example, we assumed that US people would be vaccinated on 1^st^ November, 2020 and can reach the targeted coverage within one month, based on a letter that US CDC asked governors to be prepared for COVID-19 vaccine distribution by 1^st^ November.^22^ We constructed four scenarios: (1) the social interaction returns to the pre-epidemic level within one month after Nov 1 in the absence of vaccine but the face mask use maintains its current level, i.e., no social distancing+baseline face mask use+no vaccine; (2) no social distancing+baseline face mask use+vaccine (with different effectiveness and coverage); (3) no social distancing+half of baseline face mask use+vaccine; (4) no social distancing+no face mask use+vaccine. It was assumed that natural and vaccine-induced immunity would last at least one year. Based on the estimated parameters, we simulate the epidemic trend of daily confirmed cases and deaths from November 1, 2020 to November 1, 2021.

We calculated the number of averted infections and deaths after one-year vaccination for scenarios (2)-(4), compared with scenario (1) (**Tables S5-S8**), and plotted them as a function of vaccine effectiveness and coverage (**Figures 3-4**). In these results, we define the threshold of vaccination curve as the combination of vaccine effectiveness and coverage such that social distancing restrictions may be relaxed while the COVID-19 epidemic can be retained at a very low endemic level or eliminated. We also performed a similar plot for later vaccination initiating time in 1^st^ December, 2020 (**Figures S2-S3**) and 1^st^ January, 2021 (**Figures S4-S5**).

**New York**

In the State of New York, if the social distancing was relaxed to the pre-epidemic level while maintaining the current face mask use without vaccine, the estimated number of COVID-19 infections and deaths within one year would be 2.71 (95% CIs: 2.55-2.87) million and 222,056 (201,188-242,924).

If the current face mask use rate was maintained and introducing the weak vaccine with effectiveness 50% (**Figure 3a, 4a**), then 50% coverage could avert 2.49 (2.37-2.61) million infections and 203,445 (189,366-217,525) deaths, 75% coverage could avert 2.65 (2.51-2.79) million infections and 216,290 (197,944-234,635) deaths, and 100% coverage could avert 2.68 (2.53-2.83) million infections and 218,854 (199,337-238,372) deaths. If the moderate vaccine effectiveness was 80%, then 50%, 75%, 100% coverage could avert 2.66 (2.51-2.80) million, 2.68 (2.53-2.84) million, 2.69 (2.54-2.85) million infections, and 216,698 (198,143-235,252), 218,970 (199,382-238,557), 219,956 (199,943-239,969) deaths, respectively. If the strong vaccine effectiveness was 100%, then 50%, 75%, 100% coverage could avert 2.67 (2.52-2.82) million, 2.69 (2.53-2.84) million, 2.70 (2.54-2.86) million infections, and 218,150 (198,929-237,372), 219,466 (199,659-239,272), 220,217 (200,092-240,342) deaths, respectively.

If the face mask use decreased by 50% (**Figure 3b, 4b**), the threshold of vaccination curve showed that if the vaccine effectiveness was weak or moderate, the coverage should be greater than 77.1% or 38.0%, respectively, to suppress the pandemic. Improving vaccine coverage to 100% could avert 1.03 (0.97-1.09) million infections and 85,878 (79,831-91,924) deaths, 2.69 (2.54-2.85) million infections and 219,621 (199,756-239,485) deaths, respectively, for weak and moderate vaccine. If the vaccine effectiveness was 100%, the vaccine coverage should be greater than 24.4% and improving coverage to 100% could avert 2.70 (2.54-2.86) million infections and 220,205 (200,084-240,325) deaths.

If no face mask was used and the vaccine effectiveness was 50% (**Figure 3c, 4c**), even 100% coverage cann’t suppress the pandemic. If the vaccine effectiveness was 80%, the vaccine coverage should be greater than 55.4% and improving coverage to 100% could avert 2.69 (2.53-2.83) million infections and 218,593 (199,201-237,984) deaths. If the vaccine effectiveness was 100%, the vaccine coverage should be greater than 33.2% and improving coverage to 100% could avert 2.70 (2.54-2.86) million infections and 220,191 (200,076-240,307) deaths.

If deferring the rollout of vaccine by one-month (two-month) with 50% of the current face mask use as shown in **Figures S2-S3** (**Figures S4-S5**), the required coverage should be 76.4%, 37.9%, 24.5% (75.3%, 37.7%, 24.5%) when the vaccine effectiveness was 50%, 80%, 100%, respectively. These coverages become 55.2%, 33.2% (54.8%,33.2%) in the absence of face mask use if deferring vaccination for one-month (two-month) when the vaccine effectiveness was 80%, 100%, respectively.

**Texas**

In the State of Texas, if the social distancing was relaxed to the pre-epidemic level while maintaining the current face mask use without vaccine, the estimated number of COVID-19 infections and deaths within one year would be 3.44 (2.94-3.93) million and 74,792 (63,212-86,373).

If the current face mask use rate was maintained and introducing the vaccine with effectiveness 50% (**Figure 3d, 4d**), 50% coverage could avert 3.36 (2.87-3.85) million infections and 72,732 (61,309-84,156) deaths, 75% coverage could avert 3.41 (2.91-3.91) million infections and 73,719 (62,088-85,349) deaths, 100% coverage could avert 3.42 (2.92-3.92) million infections and 73,984 (62,340-85,628) deaths. If the vaccine effectiveness was 80%, then 50%, 75%, 100% coverage could avert 3.41 (2.91-3.91) million, 3.42 (2.92-3.92) million, 3.43 (2.93-3.92) million infections, and 73,754 (62,121-85,387), 73,999 (62,352-85,646), 74,127 (62,484-85,770) deaths, respectively. If the vaccine effectiveness was 100%, then 50%, 75%, 100% coverage could avert 3.417 (2.92-3.91) million, 3.424 (2.93-3.92) million, 3.43 (2.93-3.92) million infections, and 73,905 (62,260-85,550), 74,064 (62,418-85,709), 74,166 (62,525-85,807) deaths, respectively.

If the face mask use decreased by 50% (**Figure 3e, 4e**), the threshold of vaccination curve showed that if the vaccine effectiveness was weak or moderate, the coverage should be greater than 74.6% or 41.1%, respectively, to suppress the pandemic. Improving vaccine coverage to 100% could avert 2.77 (2.37-3.17) million infections and 60,599 (51,679-69,519) deaths, and 3.425 (2.929-3.922) million infections and 74,088 (62,444-85,732) deaths, respectively, for weak and moderate vaccine. If the vaccine effectiveness was 100%, the vaccine coverage should be greater than 29.6% and improving coverage to 100% could avert 3.429 (2.933-3.925) million infections and 74,164 (62,523-85,805) deaths.

If no face mask was used and the vaccine effectiveness was 50% (**Figure 3f, 4f**), even 100% coverage cann’t suppress the pandemic. If the vaccine effectiveness was 80%, the vaccine coverage should be greater than 56.5% and improving coverage to 100% could avert 3.422 (2.925-3.918) million infections and 74,007 (62,365-85,648) deaths. If the vaccine effectiveness was 100%, the vaccine coverage should be greater than 39.0% and improving coverage to 100% could avert 3.429 (2.933-3.924) million infections and 74,162 (62,521-85,803) deaths.

If deferring the rollout of vaccine by one-month (two-month) with 50% of the current face mask use, the required coverage should be 72.0%, 40.6%, 29.4% (67.9%, 39.5%, 28.7%) when the vaccine effectiveness was 50%, 80%, 100%, respectively. These coverages become 56.1%, 38.9% (55.2%,38.6%) in the absence of face mask use if deferring vaccination for one-month (two-month) when the vaccine effectiveness was 80%, 100%, respectively.

**Florida**

In the State of Florida, if the social distancing was relaxed to the pre-epidemic level while maintaining the current face mask use without vaccine, the estimated number of COVID-19 infections and deaths within one year would be 2.08 (1.62-2.54) million and 57,540 (44,177-70,902).

If the current face mask use rate was maintained and introducing the vaccine with effectiveness 50% (**Figure 3g, 4g**), 50% coverage could avert 2.01 (1.54-2.48) million infections and 55,001 (41,197-68,805) deaths, 75% coverage could avert 2.04 (1.58-2.51) million infections and 56,034 (42,216-69,852) deaths, 100% coverage could avert 2.06 (1.59-2.52) million infections and 56,406 (42,661-70,152) deaths. If the vaccine effectiveness was 80%, then 50%, 75%, 100% coverage could avert 2.05 (1.58-2.51) million, 2.06 (1.59-2.52) million, 2.06 (1.60-2.53) million infections, and 56,065 (42,247-69,882), 56,402 (42,652-70,153), 56,610 (42,918-70,301) deaths, respectively. If the vaccine effectiveness was 100%, then 50%, 75%, 100% coverage could avert 2.05 (1.59-2.52) million, 2.061 (1.597-2.524) million, 2.067 (1.605-2.528) million infections, and 56,260 (42,476-70,044), 56,496 (42,770-70,221), 56,668 (42,995-70,342) deaths, respectively.

If the face mask use decreased by 50% (**Figure 3h, 4h**), the threshold of vaccination curve showed that if the vaccine effectiveness was weak or moderate, the coverage should be greater than 55.0% or 32.2%, respectively, to suppress the pandemic. Improving vaccine coverage to 100% could avert 2.02 (1.56-2.49) million infections and 55,500 (41,752-69,249) deaths, and 2.064 (1.601-2.526) million infections and 56,582 (42,883-70,281) deaths, respectively, for weak and moderate vaccine. If the vaccine effectiveness was 100%, the vaccine coverage should be greater than 23.4% and improving coverage to 100% could avert 2.067 (1.605-2.528) million infections and 56,667 (42,993-70,340) deaths.

If no face mask was used and the vaccine effectiveness was 50% (**Figure 3i, 4i**), the vaccine coverage should be greater than 87.8% and improving coverage to 100% could avert 725,984 (554,592-897,375) infections and 20,307 (15,142-25,473) deaths. If the vaccine effectiveness was 80%, the vaccine coverage should be greater than 47.8% and improving coverage to 100% could avert 2.062 (1.599-2.525) million infections and 56,542 (42,832-70,251) deaths. If the vaccine effectiveness was 100%, the vaccine coverage should be greater than 34.3% and improving coverage to 100% could avert 2.067 (1.605-2.528) million infections and 56,665 (42,992-70,339) deaths.

If deferring the rollout of vaccine by one-month (two-month) with 50% of the current face mask use, the required coverage should be 53.8%, 31.7%, 23.6% (51.7%, 30.8%, 23.0%) when the vaccine effectiveness was 50%, 80%, 100%, respectively. These coverages become 47.5%, 34.2% (46.9%,34.0%) in the absence of face mask use if deferring vaccination for one-month (two-month) when the vaccine effectiveness was 80%, 100%, respectively.

**California**

In the State of California, if the social distancing was relaxed to the pre-epidemic level while maintaining the current face mask use without vaccine, the estimated number of COVID-19 infections and deaths within one year would be 1.46 (0.78-2.13) million and 29,988 (15,461-44,515).

If the current face mask use rate was maintained and introducing the vaccine with effectiveness 50% (**Figure 3j, 4j**), 50% coverage could avert 1.42 (0.761-2.088) million infections and 28,757 (14,789-42,726) deaths, 75% coverage could avert 1.436 (0.766-2.106) million infections and 29,016 (14,896-43,136) deaths, 100% coverage could avert 1.442 (0.770-2.114) million infections and 29,146 (14,959-43,332) deaths. If the vaccine effectiveness was 80%, then 50%, 75%, 100% coverage could avert 1.437 (0.767-2.106) million, 1.442 (0.769-2.114) million, 1.446 (0.772-2.119) million infections, and 29,024 (14,900-43,149), 29,143 (14,957-43,330), 29,228 (15,000-43,456) deaths, respectively. If the vaccine effectiveness was 100%, then 50%, 75%, 100% coverage could avert 1.439 (0.768-2.111) million, 1.443 (0.770-2.116) million, 1.447 (0.772-2.121) million infections, and 29,091 (14,930-43,252), 29,181 (14,975-43,386), 29,254 (15,014-43,494) deaths, respectively.

If the face mask use decreased by 50% (**Figure 3k, 4k**), the threshold of vaccination curve showed that if the vaccine effectiveness was weak or moderate, the coverage should be greater than 94.2% or 56.8%, respectively, to suppress the pandemic. Improving vaccine coverage to 100% could avert 0.91 (0.52-1.31) million infections and 18,398 (10,156-26,640) deaths, 1.444 (0.771-2.118) million infections and 29,200 (14,986-43,414) deaths, respectively, for weak and moderate vaccine. If the vaccine effectiveness was 100%, the vaccine coverage should be greater than 45.7% and improving coverage to 100% could avert 1.447 (0.772-2.121) million infections and 29,253 (15,013-43,492) deaths.

If no face mask was used and the vaccine effectiveness was 50% (**Figure 3l, 4l**), even 100% coverage cann’t suppress the pandemic. If the vaccine effectiveness was 80%, the vaccine coverage should be greater than 77.8% and improving coverage to 100% could avert 1.442 (0.769-2.114) million infections and 29,140 (14,957-43,322) deaths. If the vaccine effectiveness was 100%, the vaccine coverage should be greater than 58.0% and improving coverage to 100% could avert 1.447 (0.772-2.121) million infections and 29,251 (15,012-43,490) deaths.

If deferring the rollout of vaccine by one-month (two-month) with 50% of the current face mask use, the required coverage should be 89.6%, 54.8%, 44.4% (84.5%, 52.5%, 42.9%) when the vaccine effectiveness was 50%, 80%, 100%, respectively. These coverages become 76.6%, 58.9% (74.0%,57.5%) in the absence of face mask use if deferring vaccination for one-month (two-month) when the vaccine effectiveness was 80%, 100%, respectively.

**Table S1**. The values of parameters based on references or estimated by nonlinear least-squares (NLS) method in New York State.

| Parameter | Description | Range or 95% CI from NLS | Source |
| --- | --- | --- | --- |
| 1/*k*_1_ | The mean incubation time (days) | 5.2 (4.1-7.0) | ^2^ |
| 1$/k_{2}$ | The mean time from symptoms onset to natural recovery (days) | 10 | ^3^ |
| $r$ | The mean number of members in a family | 4 | ^4^ |
| 1$/\gamma_{1}$ | The average recovery period for diagnosed mild/moderate cases (days) | 7 | ^5,6^ |
| 1/$\gamma_{0}$ | The mean time for natural recovery (days) | 9.6620 (9.1590-10.2235) | NLS |
| 1$/\alpha_{1}$ | The average period from symptoms onset to diagnose for mild/moderate cases (days) | 6.9974 (6.4824-7.6013) | NLS |
| 1$/\alpha_{2}$ | The average diagnose period for severe/critical cases (days) | 2.0421 (2.0187-2.0660) | NLS |
| 1$/\gamma_{2}$ | The average recovery period for diagnosed severe/critical cases | $21-1/\alpha_{2}$ | ^6^ |
| $1/\xi$ | The mean recovery period for infected family members (days) | 28 | ^7^ |
| E(0) | The initial value of latent individuals | 99.9998 (99.7721-100.2276) | NLS |
| A(0) | The initial value of asymptomatic individuals | 10.0000 (9.7728-10.2272) | NLS |
| I_1_(0) | The initial value of undiagnosed mild/moderate cases | 10.0000 (9.7724-10.2277) | NLS |
| I_2_(0) | The initial value of undiagnosed severe/critical individuals | 100.0000 (99.7730-100.2271) | NLS |
| $\beta$ | The per-act transmission probability in contact with infected individuals with symptoms | 0.0305 (0.0304-0.0306) | NLS |
| $\varepsilon$ | The reduction in per-act transmission probability if infection is in latent and asymptomatic stage | 75% | ^8^ |
| $\rho$ | The probability that an individual is asymptomatic | 0.5092 (0.4978-0.5206) | NLS |
| $k_{3}$ | The progression rate from diagnosed mild/moderate stage to diagnosed severe/critical stage | 0.0123 (0.0112-0.0134)$\times k_{2}$ | NLS |
| $m_{1}(t)$ | The average number of daily contacts in the public settings (before reopening) | $m_{ini}+\frac{0.2m_{ini}-m_{ini}}{1+\exp(-m_{0}(t-t_{ini}))}$ | ^12,23^ |
|  | The average number of daily contacts in the public settings (after reopening) | ${0.2m}_{ini}+\frac{3.3812\times0.2m_{ini}-{0.2m}_{ini}}{1+\exp(-m_{0}(t-t_{reopen}))}$ | NLS |
| $m_{ini}$ | Base daily contact number in the public settings | 42.0000 (41.9886-42.0114) | NLS |
| $m_{0}$ | Change rate of daily contact number | 0.7966 (0.7852-0.8080) | NLS |
| $t_{ini}$ | The time when the contact number is half of maximal and minimal contact number in public settings (before reopening) | 28.0137 (27.4445-28.5828) | NLS |
| $t_{reopen}$ | The time when the contact number is half of maximal and minimal contact number in public settings (after reopening) | 100 | ^24^ |
| $m_{2}(t)$ | The average number of daily contacts in the households (before reopening) | $4+\frac{8-4}{1+\exp(-m_{0}(t-t_{ini}))}$ | ^13^ |
|  | The average number of daily contacts in the households (after reopening) | $8+\frac{0.8000\times8-8}{1+\exp(-m_{0}(t-t_{reopen}))}$ | NLS |
| $q(t)$ | The usage percentage of handwashing | $q_{ini}+\frac{\bar{q}-q_{ini}}{1+\exp(-m_{0}(t-t_{ini}))}$ | ^14^ |
| $q_{ini}$ | Base percentage of handwashing before the epidemic | 77% | ^14^ |
| $\bar{q}$ | Maximal percentage of handwashing during the epidemic | 95% | ^15^ |
| $\theta_{2}$ | The effectiveness of handwashing in preventing infection | 0.42 (0.1-0.95) | ^16^ |
| $p_{1}(t)$ | The usage percentage of face mask in the public settings | $p_{ini}+\frac{\bar{p}-p_{ini}}{1+\exp(-(t-t_{p}))}$ | ^25^ |
| $p_{ini}$ | Base percentage of face mask usage in the public settings before the Executive Order on face mask use | 0% | Asumed |
| $\bar{p}$ | Percentage of face mask usage in the public settings after the Executive Order on face mask use | 76.6% | ^18,19^ |
| $t_{p}$ | The time when face mask usage in the public settings is half of the maximal face mask usage rate | 42 | ^17^ |
| $p_{2}$ | The usage percentage of mask in the households | 0% | ^25^ |
| $\theta_{1}$ | The effectiveness of mask in preventing infection | 0.85 (0.66-0.93) | ^21^ |
| $\mu_{1}$ | Disease-induced death rate of undiagnosed severe/critical cases | 0.0500 (0.0489-0.0511) | NLS |
| $\mu_{2}$ | Disease-induced death rate of diagnosed severe/critical cases | ${0.1131 (0.1017-0.1245)\times\mu}_{1}$ | NLS |

**Table S2**. The values of parameters based on references or estimated by nonlinear least-squares (NLS) method in Texas State. The other parameters are the same as **Table S1**.

| Parameter | Description | Range or 95% CI from NLS | Source |
| --- | --- | --- | --- |
| 1/$\gamma_{0}$ | The mean time for natural recovery (days) | 9.2849 (8.3978-10.3817) | NLS |
| 1$/\alpha_{1}$ | The average period from symptoms onset to diagnose for mild/moderate cases (days) | 5.6369 (5.4618-5.8236) | NLS |
| 1$/\alpha_{2}$ | The average diagnose period for severe/critical cases (days) | 2.6760 (2.7600-2.5969) | NLS |
| E(0) | The initial value of latent individuals | 1.5036 (1.3898-1.6173) | NLS |
| A(0) | The initial value of asymptomatic individuals | 7.5985 (7.3713-7.8257) | NLS |
| I_1_(0) | The initial value of undiagnosed mild/moderate cases | 1.3482 (1.2344-1.4620) | NLS |
| I_2_(0) | The initial value of undiagnosed severe/critical individuals | 2.2276 (2.0005-2.4548) | NLS |
| $\beta$ | The per-act transmission probability in contact with infected individuals with symptoms | 0.0303 (0.0302-0.0304) | NLS |
| $\rho$ | The probability that an individual is asymptomatic | 0.4958 (0.4844-0.5072) | NLS |
| $k_{3}$ | The progression rate from diagnosed mild/moderate stage to diagnosed severe/critical stage | 0.0507 (0.0393-0.0621)$\times k_{2}$ | NLS |
| $m_{1}(t)$ | The average number of daily contacts in the public settings (before reopening) | $m_{ini}+\frac{0.2m_{ini}-m_{ini}}{1+\exp(-m_{0}(t-t_{ini}))}$ | ^12,23^ |
|  | The average number of daily contacts in the public settings (after reopening) | ${0.2m}_{ini}+\frac{2.2636\times0.2m_{ini}-{0.2m}_{ini}}{1+\exp(-m_{0}(t-t_{reopen}))}$ | NLS |
| $m_{ini}$ | Base daily contact number in the public settings | 37.0665 (37.0654-37.0676) | NLS |
| $m_{0}$ | Change rate of daily contact number | 0.2084 (0.1970-0.2198) | NLS |
| $t_{ini}$ | The time when the contact number is half of maximal and minimal contact number in public settings (before reopening) | 32.9861 (31.8491-34.1230) | NLS |
| $t_{reopen}$ | The time when the contact number is half of maximal and minimal contact number in public settings (after reopening) | 79 | ^26^ |
| $m_{2}(t)$ | The average number of daily contacts in the households (before reopening) | $4+\frac{8-4}{1+\exp(-m_{0}(t-t_{ini}))}$ | ^13^ |
|  | The average number of daily contacts in the households (after reopening) | $8+\frac{0.7585\times8-8}{1+\exp(-m_{0}(t-t_{reopen}))}$ | NLS |
| $p_{1}(t)$ | The usage percentage of face mask in the public settings | $p_{ini}+\frac{\bar{p}-p_{ini}}{1+\exp(-0.1(t-t_{p}))}$ | ^25^ |
| $\bar{p}$ | Percentage of face mask usage in the public settings after the Executive Order on face mask use | 71.7% | ^18,19^ |
| $t_{p}$ | The time when face mask usage in the public settings is half of the maximal face mask usage rate | 149 | ^27^ |
| $\mu_{1}$ | Disease-induced death rate of undiagnosed severe/critical cases | 0.0096 (0.0094-0.0098) | NLS |
| $\mu_{2}$ | Disease-induced death rate of diagnosed severe/critical cases | ${0.1981 (0.1867-0.2095)\times\mu}_{1}$ | NLS |

**Table S3**. The values of parameters based on references or estimated by nonlinear least-squares (NLS) method in Florida State. The other parameters are the same as **Table S1**.

| Parameter | Description | Range or 95% CI from NLS | Source |
| --- | --- | --- | --- |
| 1/$\gamma_{0}$ | The mean time for natural recovery (days) | 6.9982 (6.4813-7.6047) | NLS |
| 1$/\alpha_{1}$ | The average period from symptoms onset to diagnose for mild/moderate cases (days) | 4.8780 (4.7463-5.0171) | NLS |
| 1$/\alpha_{2}$ | The average diagnose period for severe/critical cases (days) | 2.8961 (2.8038-2.9946) | NLS |
| E(0) | The initial value of latent individuals | 98.4149 (96.1388-100.6910) | NLS |
| A(0) | The initial value of asymptomatic individuals | 1.0130 (0.8994-1.1265) | NLS |
| I_1_(0) | The initial value of undiagnosed mild/moderate cases | 1.0000 (0.8861-1.1140) | NLS |
| I_2_(0) | The initial value of undiagnosed severe/critical individuals | 98.7749 (96.2714-101.2785) | NLS |
| $\beta$ | The per-act transmission probability in contact with infected individuals with symptoms | 0.0300 (0.0297-0.0303) | NLS |
| $\rho$ | The probability that an individual is asymptomatic | 0.5000 (0.4886-0.5114) | NLS |
| $k_{3}$ | The progression rate from diagnosed mild/moderate stage to diagnosed severe/critical stage | 0.0497 (0.0383-0.0611)$\times k_{2}$ | NLS |
| $m_{1}(t)$ | The average number of daily contacts in the public settings (before reopening) | $m_{ini}+\frac{0.2m_{ini}-m_{ini}}{1+\exp(-m_{0}(t-t_{ini}))}$ | ^12,23^ |
|  | The average number of daily contacts in the public settings (after reopening) | ${0.2m}_{ini}+\frac{2.8500\times0.2m_{ini}-{0.2m}_{ini}}{1+\exp(-m_{0}(t-t_{reopen}))}$ | NLS |
| $m_{ini}$ | Base daily contact number in the public settings | 34.0000 (33.8861-34.1138) | NLS |
| $m_{0}$ | Change rate of daily contact number | 0.1000 (0.0886-0.1114) | NLS |
| $t_{ini}$ | The time when the contact number is half of maximal and minimal contact number in public settings (before reopening) | 31.3877 (30.2508-32.5246) | NLS |
| $t_{reopen}$ | The time when the contact number is half of maximal and minimal contact number in public settings (after reopening) | 71 | ^28^ |
| $m_{2}(t)$ | The average number of daily contacts in the households (before reopening) | $4+\frac{8-4}{1+\exp(-m_{0}(t-t_{ini}))}$ | ^13^ |
|  | The average number of daily contacts in the households (after reopening) | $8+\frac{0.7500\times8-8}{1+\exp(-m_{0}(t-t_{reopen}))}$ | NLS |
| $p_{1}(t)$ | The usage percentage of face mask in the public settings | $p_{ini}+\frac{\bar{p}-p_{ini}}{1+\exp(-0.5(t-t_{p}))}$ | ^25^ |
| $\bar{p}$ | Percentage of face mask usage in the public settings after the Executive Order on face mask use | 58.7% | ^18,19^ |
| $t_{p}$ | The time when face mask usage in the public settings is half of the maximal face mask usage rate | 124 | ^29^ |
| $\mu_{1}$ | Disease-induced death rate of undiagnosed severe/critical cases | 0.0145 (0.0134-0.0156) | NLS |
| $\mu_{2}$ | Disease-induced death rate of diagnosed severe/critical cases | ${0.1600 (0.1486-0.1714)\times\mu}_{1}$ | NLS |

**Table S4**. The values of parameters based on references or estimated by nonlinear least-squares (NLS) method in California State. The other parameters are the same as **Table S1**.

| Parameter | Description | Range or 95% CI from NLS | Source |
| --- | --- | --- | --- |
| 1/$\gamma_{0}$ | The mean time for natural recovery (days) | 7.0182 (6.5006-7.6253) | NLS |
| 1$/\alpha_{1}$ | The average period from symptoms onset to diagnose for mild/moderate cases (days) | 6.3013 (5.8805-6.7870) | NLS |
| 1$/\alpha_{2}$ | The average diagnose period for severe/critical cases (days) | 2.1309 (2.0805-2.1837) | NLS |
| E(0) | The initial value of latent individuals | 1.3844 (1.2705-1.4982) | NLS |
| A(0) | The initial value of asymptomatic individuals | 17.9226 (17.1272-18.7180) | NLS |
| I_1_(0) | The initial value of undiagnosed mild/moderate cases | 1.0285 (0.9148-1.1422) | NLS |
| I_2_(0) | The initial value of undiagnosed severe/critical individuals | 1.7759 (1.5484-2.0034) | NLS |
| $\beta$ | The per-act transmission probability in contact with infected individuals with symptoms | 0.0325 (0.0324-0.0326) | NLS |
| $\rho$ | The probability that an individual is asymptomatic | 0.5000 (0.4886-0.5114) | NLS |
| $k_{3}$ | The progression rate from diagnosed mild/moderate stage to diagnosed severe/critical stage | 0.0059 (0.0058-0.0060)$\times k_{2}$ | NLS |
| $m_{1}(t)$ | The average number of daily contacts in the public settings (before reopening) | $m_{ini}+\frac{0.2m_{ini}-m_{ini}}{1+\exp(-m_{0}(t-t_{ini}))}$ | ^12,23^ |
|  | The average number of daily contacts in the public settings (after reopening) | ${0.2m}_{ini}+\frac{2.5002\times0.2m_{ini}-{0.2m}_{ini}}{1+\exp(-m_{0}(t-t_{reopen}))}$ | NLS |
| $m_{ini}$ | Base daily contact number in the public settings | 39.0000 (38.9431-39.0569) | NLS |
| $m_{0}$ | Change rate of daily contact number | 0.2066 (0.1895-0.2237) | NLS |
| $t_{ini}$ | The time when the contact number is half of maximal and minimal contact number in public settings (before reopening) | 30.0002 (28.8616-31.1388) | NLS |
| $t_{reopen}$ | The time when the contact number is half of maximal and minimal contact number in public settings (after reopening) | 117 | ^30^ |
| $m_{2}(t)$ | The average number of daily contacts in the households (before reopening) | $4+\frac{8-4}{1+\exp(-m_{0}(t-t_{ini}))}$ | ^13^ |
|  | The average number of daily contacts in the households (after reopening) | $8+\frac{0.7500\times8-8}{1+\exp(-m_{0}(t-t_{reopen}))}$ | NLS |
| $p_{1}(t)$ | The usage percentage of face mask in the public settings | $p_{ini}+\frac{\bar{p}-p_{ini}}{1+\exp(-0.1(t-t_{p}))}$ | ^25^ |
| $\bar{p}$ | Percentage of face mask usage in the public settings after the Executive Order on face mask use | 76.6% | ^18,19^ |
| $t_{p}$ | The time when face mask usage in the public settings is half of the maximal face mask usage rate | 72 | []^31^ |
| $\mu_{1}$ | Disease-induced death rate of undiagnosed severe/critical cases | 0.0074 (0.0072-0.0076) | NLS |
| $\mu_{2}$ | Disease-induced death rate of diagnosed severe/critical cases | ${0.3060 (0.2946-0.3173)\times\mu}_{1}$ | NLS |

**Table S5**. If the social distancing was relaxed to the pre-epidemic level while maintaining the current face mask use without vaccine, the estimated number of COVID-19 infections and deaths within one year in New York state were 2.71 (95% CIs: 2.55-2.87) million and 222,056 (201,188-242,924). The averted infections and deaths for different vaccine effectiveness and coverage are listed as follows.

| Simulation scenarios (New York State) | | Averted infections | Averted deaths |
| --- | --- | --- | --- |
| No social distancing  + baseline face mask coverage (76.6%) | Effectiveness=50%,  Coverage=50% | 2.49 (2.37-2.61) million | 203,445 (189,366-217,525) |
|  | Effectiveness=50%,  Coverage=75% | 2.65 (2.51-2.79) million | 216,290 (197,944-234,635) |
|  | Effectiveness=50%,  Coverage=100% | 2.68 (2.53-2.83) million | 218,854 (199,337-238,372) |
|  | Effectiveness=80%,  Coverage=50% | 2.66 (2.51-2.80) million | 216,698 (198,143-235,252) |
|  | Effectiveness=80%,  Coverage=75% | 2.68 (2.53-2.84) million | 218,970 (199,382-238,557) |
|  | Effectiveness=80%,  Coverage=100% | 2.69 (2.54-2.85) million | 219,956 (199,943-239,969) |
|  | Effectiveness=100%,  Coverage=50% | 2.67 (2.52-2.82) million | 218,150 (198,929-237,372) |
|  | Effectiveness=100%,  Coverage=75% | 2.69 (2.53-2.84) million | 219,466 (199,659-239,272) |
|  | Effectiveness=100%,  Coverage=100% | 2.70 (2.54-2.86) million | 220,217 (200,092-240,342) |
| No social distancing  + half of baseline face mask coverage (38.3%) | Effectiveness=50%,  Coverage=100% | 1.03 (0.97-1.09) million | 85,878 (79,831-91,924) |
|  | Effectiveness=80%,  Coverage=100% | 2.69 (2.54-2.85) million | 219,621 (199,756-239,485) |
|  | Effectiveness=100%,  Coverage=100% | 2.70 (2.54-2.86) million | 220,205 (200,084-240,325) |
| No social distancing  +no face mask use | Effectiveness=80%,  Coverage=100% | 2.69 (2.53-2.83) million | 218,593 (199,201-237,984) |
|  | Effectiveness=100%,  Coverage=100% | 2.70 (2.54-2.86) million | 220,191 (200,076-240,307) |

**Table S6**. If the social distancing was relaxed to the pre-epidemic level while maintaining the current face mask use without vaccine, the estimated number of COVID-19 infections and deaths within one year in Texas state were 3.44 (2.94-3.93) million and 74,792 (63,212-86,373). The averted infections and deaths for different vaccine effectiveness and coverage are listed as follows.

| Simulation scenarios (Texas State) | | Averted infections | Averted deaths |
| --- | --- | --- | --- |
| No social distancing  + baseline face mask coverage (71.7%) | Effectiveness=50%,  Coverage=50% | 3.36 (2.87-3.85) million | 72,732 (61,309-84,156) |
|  | Effectiveness=50%,  Coverage=75% | 3.41 (2.91-3.91) million | 73,719 (62,088-85,349) |
|  | Effectiveness=50%,  Coverage=100% | 3.42 (2.92-3.92) million | 73,984 (62,340-85,628) |
|  | Effectiveness=80%,  Coverage=50% | 3.41 (2.91-3.91) million | 73,754 (62,121-85,387) |
|  | Effectiveness=80%,  Coverage=75% | 3.42 (2.92-3.92) million | 73,999 (62,352-85,646) |
|  | Effectiveness=80%,  Coverage=100% | 3.43 (2.93-3.92) million | 74,127 (62,484-85,770) |
|  | Effectiveness=100%,  Coverage=50% | 3.417 (2.92-3.91) million | 73,905 (62,260-85,550) |
|  | Effectiveness=100%,  Coverage=75% | 3.424 (2.93-3.92) million | 74,064 (62,418-85,709) |
|  | Effectiveness=100%,  Coverage=100% | 3.43 (2.93-3.92) million | 74,166 (62,525-85,807) |
| No social distancing  + half of baseline face mask coverage (35.9%) | Effectiveness=50%,  Coverage=100% | 2.77 (2.37-3.17) million | 60,599 (51,679-69,519) |
|  | Effectiveness=80%,  Coverage=100% | 3.425 (2.929-3.922) million | 74,088 (62,444-85,732) |
|  | Effectiveness=100%,  Coverage=100% | 3.429 (2.933-3.925) million | 74,164 (62,523-85,805) |
| No social distancing  +no face mask use | Effectiveness=80%,  Coverage=100% | 3.422 (2.925-3.918) million | 74,007 (62,365-85,648) |
|  | Effectiveness=100%,  Coverage=100% | 3.429 (2.933-3.924) million | 74,162 (62,521-85,803) |

**Table S7**. If the social distancing was relaxed to the pre-epidemic level while maintaining the current face mask use without vaccine, the estimated number of COVID-19 infections and deaths within one year in Florida state were 2.08 (1.62-2.54) million and 57,540 (44,177-70,902). The averted infections and deaths for different vaccine effectiveness and coverage are listed as follows.

| Simulation scenarios (Florida State) | | Averted infections | Averted deaths |
| --- | --- | --- | --- |
| No social distancing  + baseline face mask coverage (58.7%) | Effectiveness=50%,  Coverage=50% | 2.01 (1.54-2.48) million | 55,001 (41,197-68,805) |
|  | Effectiveness=50%,  Coverage=75% | 2.04 (1.58-2.51) million | 56,034 (42,216-69,852) |
|  | Effectiveness=50%,  Coverage=100% | 2.06 (1.59-2.52) million | 56,406 (42,661-70,152) |
|  | Effectiveness=80%,  Coverage=50% | 2.05 (1.58-2.51) million | 56,065 (42,247-69,882) |
|  | Effectiveness=80%,  Coverage=75% | 2.06 (1.59-2.52) million | 56,402 (42,652-70,153) |
|  | Effectiveness=80%,  Coverage=100% | 2.06 (1.60-2.53) million | 56,610 (42,918-70,301) |
|  | Effectiveness=100%,  Coverage=50% | 2.05 (1.59-2.52) million | 56,260 (42,476-70,044) |
|  | Effectiveness=100%,  Coverage=75% | 2.061 (1.597-2.524) million | 56,496 (42,770-70,221) |
|  | Effectiveness=100%,  Coverage=100% | 2.067 (1.605-2.528) million | 56,668 (42,995-70,342) |
| No social distancing  + half of baseline face mask coverage (29.4%) | Effectiveness=50%,  Coverage=100% | 2.02 (1.56-2.49) million | 55,500 (41,752-69,249) |
|  | Effectiveness=80%,  Coverage=100% | 2.064 (1.601-2.526) million | 56,582 (42,883-70,281) |
|  | Effectiveness=100%,  Coverage=100% | 2.067 (1.605-2.528) million | 56,667 (42,993-70,340) |
| No social distancing  +no face mask use | Effectiveness=80%,  Coverage=100% | 2.062 (1.599-2.525) million | 56,542 (42,832-70,251) |
|  | Effectiveness=100%,  Coverage=100% | 2.067 (1.605-2.528) million | 56,665 (42,992-70,339) |

**Table S8**. If the social distancing was relaxed to the pre-epidemic level while maintaining the current face mask use without vaccine, the estimated number of COVID-19 infections and deaths within one year in California state were 1.46 (0.78-2.13) million and 29,988 (15,461-44,515). The averted infections and deaths for different vaccine effectiveness and coverage are listed as follows.

| Simulation scenarios (California State) | | Averted infections | Averted deaths |
| --- | --- | --- | --- |
| No social distancing  + baseline face mask coverage (76.6%) | Effectiveness=50%,  Coverage=50% | 1.42 (0.761-2.088) million | 28,757 (14,789-42,726) |
|  | Effectiveness=50%,  Coverage=75% | 1.436 (0.766-2.106) million | 29,016 (14,896-43,136) |
|  | Effectiveness=50%,  Coverage=100% | 1.442 (0.770-2.114) million | 29,146 (14,959-43,332) |
|  | Effectiveness=80%,  Coverage=50% | 1.437 (0.767-2.106) million | 29,024 (14,900-43,149) |
|  | Effectiveness=80%,  Coverage=75% | 1.442 (0.769-2.114) million | 29,143 (14,957-43,330) |
|  | Effectiveness=80%,  Coverage=100% | 1.446 (0.772-2.119) million | 29,228 (15,000-43,456) |
|  | Effectiveness=100%,  Coverage=50% | 1.439 (0.768-2.111) million | 29,091 (14,930-43,252) |
|  | Effectiveness=100%,  Coverage=75% | 1.443 (0.770-2.116) million | 29,181 (14,975-43,386) |
|  | Effectiveness=100%,  Coverage=100% | 1.447 (0.772-2.121) million | 29,254 (15,014-43,494) |
| No social distancing  + half of baseline face mask coverage (38.3%) | Effectiveness=50%,  Coverage=100% | 0.91 (0.52-1.31) million | 18,398 (10,156-26,640) |
|  | Effectiveness=80%,  Coverage=100% | 1.444 (0.771-2.118) million | 29,200 (14,986-43,414) |
|  | Effectiveness=100%,  Coverage=100% | 1.447 (0.772-2.121) million | 29,253 (15,013-43,492) |
| No social distancing  +no face mask use | Effectiveness=80%,  Coverage=100% | 1.442 (0.769-2.114) million | 29,140 (14,957-43,322) |
|  | Effectiveness=100%,  Coverage=100% | 1.447 (0.772-2.121) million | 29,251 (15,012-43,490) |

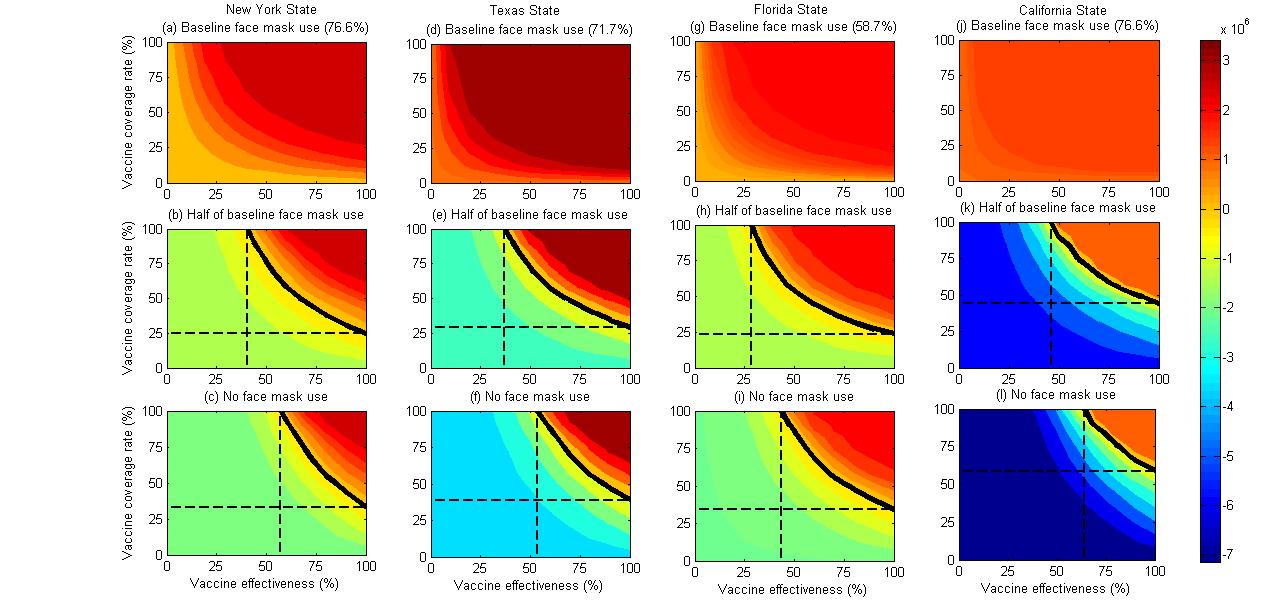

**Figure S2.** Contour plots of averted infections as a function of vaccine effectiveness and vaccine coverage rate in the four states when relaxing social distancing to pre-pandemic level and maintaining face mask use at the baseline level (the first line), half of the baseline level (the second line), and no use (the third line). The black solid isoclines indicate the threshold that the number of averted infections is zero. The black dashed lines correspond to the minimal vaccine effectiveness and vaccine coverage rate when the number of averted infections is zero. Vaccination was assumed to be initiated on 1^st^ December, 2020.

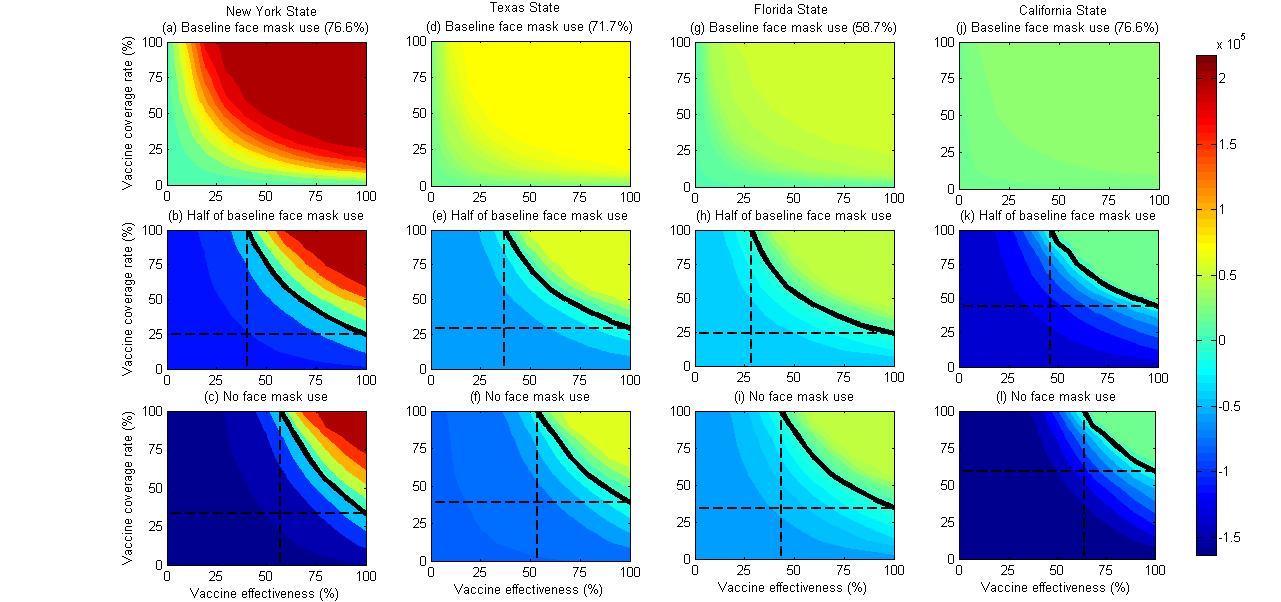

**Figure S3.** Contour plots of averted deaths as a function of vaccine effectiveness and vaccine coverage rate in the four states when relaxing social distancing to pre-pandemic level and maintaining face mask use at the baseline level (the first line), half of the baseline level (the second line), and no use (the third line). The black solid isoclines indicate the threshold that the number of averted deaths is zero. The black dashed lines correspond to the minimal vaccine effectiveness and vaccine coverage rate when the number of averted deaths is zero. Vaccination was assumed to be initiated on 1^st^ December, 2020.

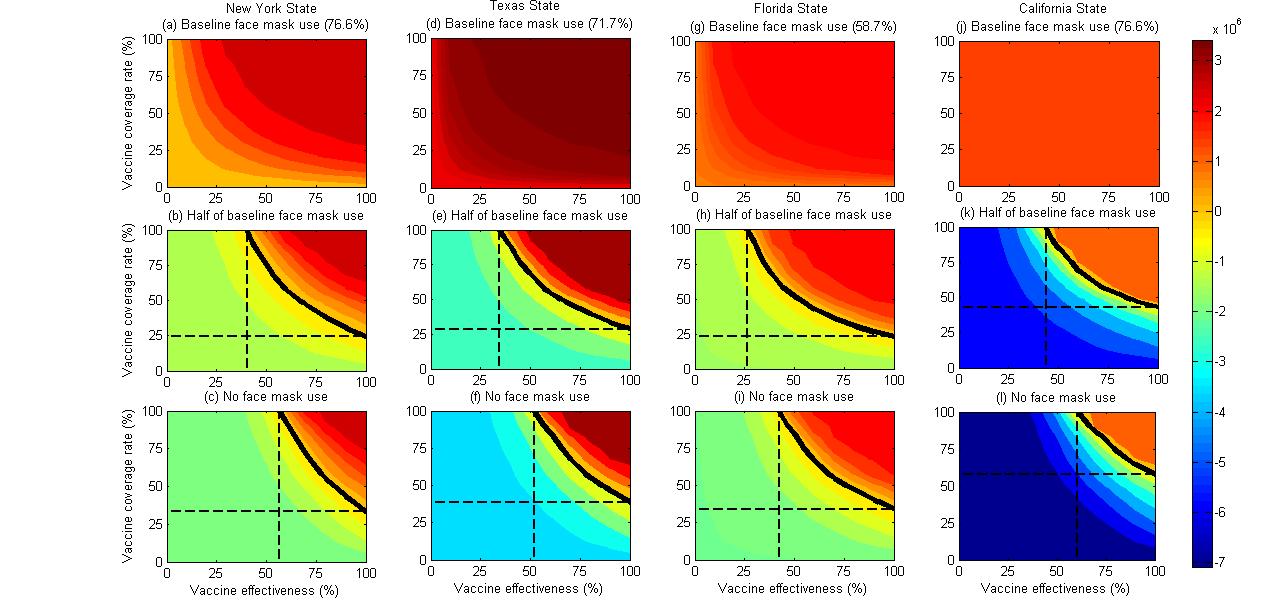

**Figure S4.** Contour plots of averted infections as a function of vaccine effectiveness and vaccine coverage rate in the four states when relaxing social distancing to pre-pandemic level and maintaining face mask use at the baseline level (the first line), half of the baseline level (the second line), and no use (the third line). The black solid isoclines indicate the threshold that the number of averted infections is zero. The black dashed lines correspond to the minimal vaccine effectiveness and vaccine coverage rate when the number of averted infections is zero. Vaccination was assumed to be initiated on 1^st^ January, 2021.

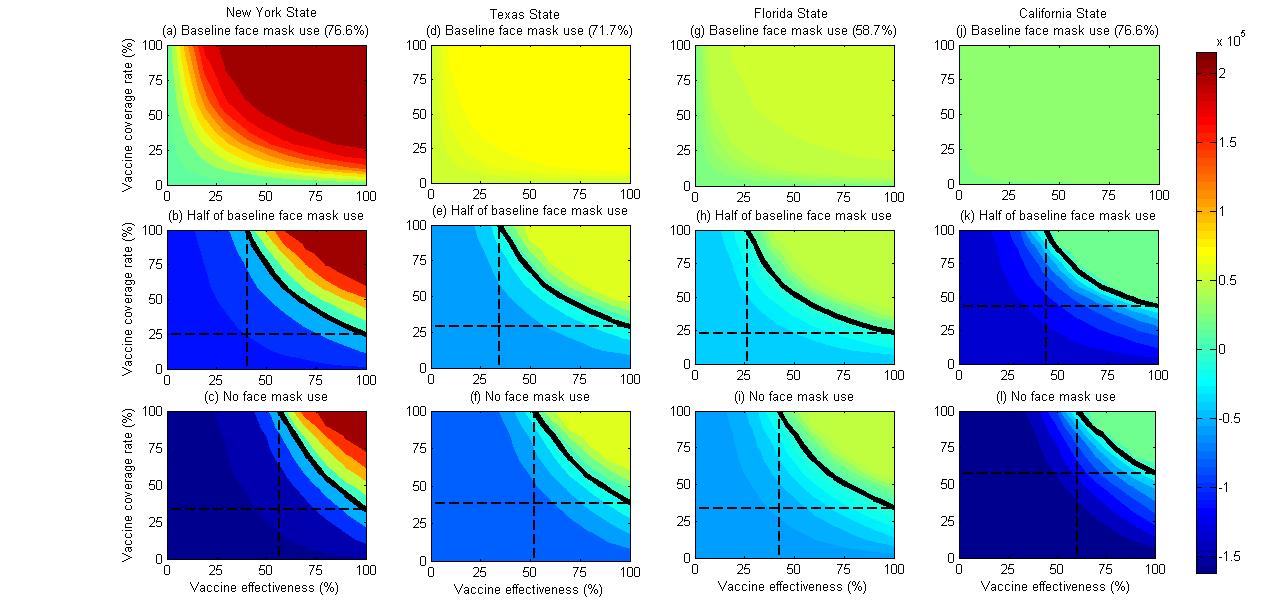

**Figure S5.** Contour plots of averted deaths as a function of vaccine effectiveness and vaccine coverage rate in four states when relaxing social distancing to pre-pandemic level and maintaining face mask use at the baseline level (the first line), half of the baseline level (the second line), and no use (the third line). The black solid isoclines indicate the threshold that the number of averted deaths is zero. The black dashed lines correspond to the minimal vaccine effectiveness and vaccine coverage rate when the number of averted deaths is zero. Vaccination was assumed to be initiated on 1^st^ January, 2021.

**References**

1.	New York City Department of Health and Mental Hygiene. *COVID-19: DATA.* Accessed April 13, 2020. https://www1.nyc.gov/site/doh/covid/covid-19-data.page

2.	Li Q, Guan X, Wu P, et al. Early transmission dynamics in Wuhan, China, of novel coronavirus–infected pneumonia. *N Engl J Med*. Published online 2020.

3.	Wang D, Hu B, Hu C, et al. Clinical characteristics of 138 hospitalized patients with 2019 novel coronavirus–infected pneumonia in Wuhan, China. *Jama*. 2020;323(11):1061–1069.

4.	Duffin E. Average size of households in the U.S. 2019. Statista. Accessed April 16, 2020. https://www.statista.com/statistics/183648/average-size-of-households-in-the-us/

5.	Huang C, Wang Y, Li X, et al. Clinical features of patients infected with 2019 novel coronavirus in Wuhan, China. *The Lancet*. 2020;395(10223):497–506.

6.	Organization WH, Organization WH. *Report of the Who-China Joint Mission on Coronavirus Disease 2019 (Covid-19)*.; 2020.

7.	Shen M, Peng Z, Guo Y, et al. Assessing the effects of metropolitan-wide quarantine on the spread of COVID-19 in public space and households. *Int J Infect Dis*. Published online 2020.

8.	Prem K, Liu Y, Russell TW, et al. The effect of control strategies to reduce social mixing on outcomes of the COVID-19 epidemic in Wuhan, China: a modelling study. *Lancet Public Health*. Published online 2020.

9.	Zhang J, Litvinova M, Liang Y, et al. Changes in contact patterns shape the dynamics of the COVID-19 outbreak in China. *Science*. Published online 2020.

10.	Jarvis CI, Van Zandvoort K, Gimma A, et al. Quantifying the impact of physical distance measures on the transmission of COVID-19 in the UK. *BMC Med*. 2020;18:1–10.

11.	UNACAST. *Social Distancing Scoreboard*.; 2020. Accessed June 7, 2020. https://www.unacast.com

12.	Ottaway A, Kelly J. It’s a Lockdown: Cuomo Puts New York on “Pause.” Published March 20, 2020. Accessed April 16, 2020. https://www.courthousenews.com/teetering-on-lockdown-cuomo-new-york-on-pause/

13.	Ferguson NM, Cummings DA, Fraser C, Cajka JC, Cooley PC, Burke DS. Strategies for mitigating an influenza pandemic. *Nature*. 2006;442(7101):448–452.

14.	Pogrebna G, Kharlamov A. The Impact of Cross-Cultural Differences in Handwashing Patterns on the COVID-19 Outbreak Magnitude. Published online 2020. doi:10.13140/RG.2.2.23764.96649

15.	Park CL, Russell BS, Fendrich M, Finkelstein-Fox L, Hutchison M, Becker J. Americans’ COVID-19 Stress, Coping, and Adherence to CDC Guidelines. *J Gen Intern Med*. Published online 2020:1.

16.	Fung IC-H, Cairncross S. Effectiveness of handwashing in preventing SARS: a review. *Trop Med Int Health*. 2006;11(11):1749–1758.

17.	Andrew S, Froio J. *These Are the States That Require You to Wear a Face Mask in Public*. CNN; 2020. https://edition.cnn.com/2020/04/20/us/states-that-require-masks-trnd/index.html

18.	The New York Times and Dynata. *Mask-Wearing Survey Data*. New York Times; 2020. https://github.com/nytimes/covid-19-data/blob/master/mask-use/mask-use-by-county.csv

19.	Robert Wood Johnson Foundation, University of Wisconsin Population Health Institute. County Health Rankings & Roadmaps. Accessed July 5, 2015. http://www.countyhealthrankings.org

20.	Offeddu V, Yung CF, Low MSF, Tam CC. Effectiveness of masks and respirators against respiratory infections in healthcare workers: a systematic review and meta-analysis. *Clin Infect Dis*. 2017;65(11):1934–1942.

21.	Chu DK, Akl EA, Duda S, et al. Physical distancing, face masks, and eye protection to prevent person-to-person transmission of SARS-CoV-2 and COVID-19: a systematic review and meta-analysis. *The Lancet*. Published online 2020.

22.	Business Insider. US CDC asks states to prepare for distribution of potential COVID-19 vaccine by November 1. Published online 2020. https://www.businessinsider.in/science/health/news/us-cdc-asks-states-to-prepare-for-distribution-of-potential-covid-19-vaccine-by-november-1/articleshow/77905321.cms

23.	Zhang L, Shen M, Ma X, et al. What is Required to Prevent a Second Major Outbreak of the Novel Coronavirus COVID-19 Upon Lifting the Metropolitan-Wide Quarantine of Wuhan City, China: A Mathematical Modelling Study. *The Innovation*. Published online 2020. doi:http://dx.doi.org/10.2139/ssrn.3555236

24.	NYC Business. *NYC Business Reopenning Guide*. The City of New York; 2020. https://www1.nyc.gov/nycbusiness/article/reopening-guide

25.	Karson K. More than half of Americans wear masks as coronavirus’ new normal takes hold: POLL. ABC News. Accessed April 16, 2020. https://abcnews.go.com/Politics/half-americans-wear-masks-coronavirus-normal-takes-hold/story?id=70073942

26.	Governor of the State of Texas. *Relating to the Safe, Strategic Reopening of Select Services as the First Step to Open Texas in Response to Tile COVID-19 Disaster*.; 2020. https://gov.texas.gov/uploads/files/press/EO-GA-16_Opening_Texas_COVID-19_FINAL_04-17-2020.pdf

27.	Statewide mask requirement in Texas now in effect. Published online 2020. https://www.fox26houston.com/news/statewide-mask-requirement-in-texas-now-in-effect

28.	CNN. *Florida Will Start to Reopen May 4, but for Now Miami-Dade and Two Other Counties Won’t Be Included.*; 2020. https://edition.cnn.com/2020/04/29/us/florida-reopening-coronavirus/index.html

29.	New4JAX. *Most Major Florida Cities Now Require Wearing Face Masks in Public.*; 2020. https://www.news4jax.com/news/local/2020/06/19/major-florida-cities-now-require-use-of-face-mask-in-public-places/

30.	NBC. *California Is Relaxing Some of Its Criteria for Reopening. Here Are the Changes.*; 2020. https://www.nbclosangeles.com/news/coronavirus/california-reopening-stay-home-order-coronavirus-covid-19-gavin-newsom/2364760/

31.	NBC news. *California Mandates Masks for Most Public Activity as Coronavirus Numbers Surge*.; 2020. https://www.nbcnews.com/news/us-news/california-mandates-masks-most-public-activity-coronavirus-numbers-surge-n1231487
